# Effectiveness of Nirmatrelvir-Ritonavir for the treatment of patients with mild to moderate COVID-19 and at high risk of hospitalization: Systematic review and meta-analyses of observational studies

**DOI:** 10.1101/2023.03.27.23287621

**Authors:** Kathiaja Miranda Souza, Gabriela Carrasco, Robin Rojas-Cortés, Mariana Michel Barbosa, Eduardo Henrique Ferreira Bambirra, José Luis Castro, Juliana Alvares-Teodoro

## Abstract

**Objective:** To assess the effectiveness of nirmatrelvir-ritonavir in the treatment of outpatients with mild to moderate COVID-19 who are at higher risk of developing severe illness, through a systematic review with meta-analyses of observational studies.

**Methods:** A systematic search was performed, in accordance with the Cochrane search methods, to identify observational studies that met the inclusion criteria. The outcomes of mortality and hospitalization were analyzed. Search was conducted on PubMed, EMBASE, and The Cochrane Library. Two reviewers independently screened references, selected the studies, extracted the data, assessed the risk of bias using ROBINS-I tool and evaluated the quality of evidence using the GRADE tool. This study followed the PRISMA reporting guideline.

**Results:** A total of 16 observational studies and 1,482,923 patients were finally included. The results of the meta-analysis showed that in comparison to standard treatment without antivirals, nirmatrelvir-ritonavir reduced the risk of death by 62% (OR= 0.38; 95% CI: 0.30-0.46; moderate certainty of evidence). In addition, a 53% reduction in the risk of hospital admission was observed (OR = 0.47; 95% CI: 0.36–0.60, with very low certainty of evidence). For the composite outcome of hospitalization and/or mortality, there was a 56% risk reduction (OR=0.44; 95% CI: 0.31-0.64, moderate certainty of evidence).

**Conclusion:** The results suggest that nirmatrelvir-ritonavir could be effective in reducing mortality and hospitalization. The results were valid in vaccinated or unvaccinated high-risk individuals with COVID-19. Data from ongoing and future trials may further advance our understanding of the effectiveness and safety of nirmatrelvir-ritonavir and help improve treatment guidelines for COVID-19.

## Introduction

Declared a pandemic by the World Health Organization (WHO) in March 2020, COVID-19 (coronavirus disease) has posed a significant challenge to healthcare professionals, managers, and health systems, due to its rapid spread, lack of treatment, severity, and unpredictable nature. As of March 7, 2023, there were 759,408,703 confirmed cases of COVID-19, including 6,866,434 deaths (1).

WHO data indicates that about 15% of mild/moderate cases progress to severe disease requiring hospitalization and respiratory support, and 5% of patients develop the critical form requiring admission to the Intensive Care Unit (ICU). The high number of cases has resulted in a massive and sudden influx of patients to emergency services, leading to large number of hospitalizations, requiring isolation, oxygen support, intubation, and invasive mechanical ventilation (2).

In Latin America, the COVID-19 pandemic has affected countries differently. Among some of these countries, the reported incidence rate ranged from 4.59% in Jamaica to 25.6% in Chile. In contrast, Peru had the highest case fatality rate (5.1%) and Chile the lowest case fatality rate (1.3%) among the countries analyzed (3).

In December 2020, the first dose of the COVID-19 vaccine was administered, and since then, 13.01 billion doses have been given worldwide, corresponding to 68.5% of the world’s population receiving at least one dose of the vaccine. In Latin America, the proportions of vaccinated individuals vary significantly between countries. While in Jamaica 28.2% of people received at least one dose, and 24.8% received the second dose, in Chile, more than 90% of the population received two doses of the COVID-19 vaccine (1).

In the context of the appearance of new variants and, in some countries, low vaccination rates, either due to unavailability or lack of adherence, the existence of medicines capable of controlling symptoms and avoiding hospitalizations and deaths is becoming increasingly under focus. In April 2022, the WHO published a new update of the “Guideline Therapeutics and COVID-19: living guideline”. In this publication, WHO made a strong recommendation in favor of nirmatrelvir-ritonavir, for patients with mild and moderate COVID-19 at high-risk of hospital admission, qualifying it as the best therapeutic option for those patients, such as unvaccinated, elderly or immunocompromised patients. The guideline development group concluded that nirmatrelvir-ritonavir represents a superior option as it may be more effective in preventing hospitalization than the alternatives compared (standard treatment, molnupiravir and remdesivir), though with important pharmacokinetic interactions, it apparently has fewer concerns than monulpiravir regarding adverse effects, and it is easier to administer than intravenous remdesivir and monoclonal antibodies (4). The Ongoing Living Systematic Review published by Pan American Health Organization (PAHO) presented the same direction of the recommendations (5).

It is noteworthy that randomized clinical trials investigating the use of nirmatrelvir-ritonavir in non-hospitalized symptomatic COVID-19 patients with a full COVID-19 vaccination schedule and/or who are at risk of progressing to severe disease have not yet been published (5,6).

Nirmatrelvir-ritonavir is a high-cost medicine, the target population is quite large, and in several countries the medicine has yet to be approved for emergency use, marketing or reimbursement into the health system due to the uncertainties and challenges related to its efficacy, further information on safety, high risk (e.g., vaccination status), cost, and resource requirements for administration.

In order to support the pharmacotherapeutic committees, health technology assessment agencies, and other decision-making bodies for the management of patients diagnosed with COVID-19 and eligible for treatment with nirmatrelvir-ritonavir in countries, a systematic review for the assessment of the effectiveness of nirmatrelvir-ritonavir was conducted.

## Materials and methods

### Search strategy

Two independent investigators conducted a thorough literature search on PubMed, EMBASE, and The Cochrane Library. Validated filters for observational studies were applied to each database to ensure relevant results. In addition, searches were conducted on Epistemonikos and ClinicalTrials to identify possible systematic reviews and primary studies not retrieved in the main databases. The search strategies developed for each platform are detailed in the Supporting Information (S1 file: Table 1) and were executed until January 4, 2023. The records obtained from the databases were imported into Mendeley® for the identification and elimination of duplicate studies. The report was based on the Preferred Reporting Items for Systematic Reviews and Meta-analysis (PRISMA) (S1 file: Table 2). This study didńt need the approval of an ethics committee since it is a secondary study (7).

**Table 1:** Characteristics of included studies.

| Study | Study design | Characteristics of included patients. | Country | Study period | Time of follow-up | Predominant SARS-CoV-variants. | Funding |
| --- | --- | --- | --- | --- | --- | --- | --- |
| Ganatra et al., 2022 (11) | retrospective cohort | Adults aged 18 or older who were vaccinated and subsequently contracted COVID-19 at least 1 month after vaccination and were not hospitalized. | United States | 1 December 2021 to 18 April 2022 | 30 days | Not reported | Not reported |
| Yip et al., 2022 (12) | retrospective cohort | Outpatient patients, regardless of vaccination status, who attended one of the selected clinics | China | 16 February 2022 to 31 March 2022 | 30 days | Omicron | None declared |
| Wai et al., 2023 (15) | retrospective cohort | Hospitalized and non-hospitalized patients aged 60 years or older or with at least one chronic disease with mild to moderate COVID-19. | China | 22 February 2022 to 15 April 2022 | 30 days | Omicron | The Tung's Foundation, Innovation and Technology Commission of Hong Kong |
| Hedvat et al., 2022 (16) | retrospective cohort | Non-hospitalized adult solid organ transplant recipients with asymptomatic, mild, or moderate COVID-19. | United States | 16 December 2021 to 19 January 2022 | 30 days | Omicron (BA.1) | Not reported |
| Dryden-Peterson et al., 2022 (17) | retrospective cohort | Outpatient patients aged 50 years or older with COVID-19. | United States | 1 January 2022 to 17 July 2022 to | 14 days<br>28 days | Omicron (BA.1.1, BA.2, BA.2.12.1 y BA.5) | U.S. National Institutes of Health. |
| Wong et al., 2022 (18) | retrospective cohort | Outpatient patients with mild clinical presentation of COVID-19 and high risk of disease severity. | China | 26 February 2022 to 26 June 2022 | 28 days | Omicron (BA.2.2) | Health and Medical Research Fund |
| Arbel et. al, 2022 (19) | retrospective cohort | Outpatient patients aged 40 years or older with mild clinical presentation of COVID-19 and high risk of disease severity. | Israel | 9 January 2022 to 31 March 2022 | 35 days | Omicron | None declared |
| Schwartz et. al., 2023 (14) | retrospective cohort | Outpatient patients aged 18 years or older with COVID-19. | Canada | 4 April 2022 to 31 August 2022. | 30 days | Omicron | Ontario Ministry of Health (MOH); the Ministry of Long-Term Care (MLTC); Public Health Ontario |
| Aggarwal et al., 2023 (22) | retrospective cohort | All non-hospitalized patients within the Colorado healthcare system with a positive test result for SARS-CoV-2. | United States | 26 March 2022 to 25 August 2022 | 28 days | Omicron (BA.2/BA.2.12.1) | U.S. National Institutes of Health |
| Najjar-Debbiny et al., 2022 (20) | retrospective cohort | Patients ≥18 years old with COVID-19 who are not hospitalized and have at least one comorbidity or condition associated with high risk of severe COVID-19. | Israel | 1 January 2022 to 28 February 2022 | 28 days | Omicron (BA.1) | Not reported |
| Qian et al., 2022 (13) | retrospective cohort | Patients ≥18 years old with COVID-19 and a diagnosis of systemic autoimmune rheumatic disease. | United States | 23 January 2022 to 30 May 2022 | 30 days | Omicron | Rheumatology Research Foundation |
| Shah et al., 2022 (21) | retrospective cohort | Patients aged ≥18 years with COVID-19 who are not hospitalized and have at least 1 comorbidity or condition associated with a high risk of severe COVID-19. | United States | 1 April 2022 to 31 August 2022 | 30 days | Omicron | Not reported |
| Bajema et. al., 2022 (23)* | retrospective cohort | Non-hospitalized veteran patients with at least one risk factor, clinical presentation of COVID-19, and high risk of disease severity. | United States | 1 January 2022 to 28 February 2022 | 30 days<br>31 – 180 days | Omicron (B.1.1.529 y BA1.1) | Veterans Health Administration Health Services Research & Development (HSR&D) |
| Lewnard et. al., 2023 (26)* | retrospective cohort | Outpatient patients aged 12 years and older with COVID-19 within Kaiser Permanente, Southern California healthcare system. | United States | 8 April 2022 to 7 October 2022 | 30 days<br>60 days | Omicron (BA.2; BA.4 y BA.5) | US Centers for Disease Control & Prevention<br>National Institute for Allergy and Infectious Diseases of the US<br>National Institutes of Health. |
| Zhou et. al., 2022 (27)* | retrospective cohort | Outpatient patients aged 12 years and older with COVID-19 within Optum repository, with >700 hospitals and 7000 clinics from all states in the US. | United States | 22 december 2021 to 8 May 2022 | 15 days<br>30 days | Omicron | Pfizer Inc. |
| Patel et al., 2022 (24)* | retrospective cohort | Outpatient patients, aged $\geq 12$ years at study initiation, and diagnosed with COVID-19 | England | 1 december 2021 to 31 May 2022 | 28 days | Omicron (BA.1, BA.2 y BA.5) | GlaxoSmithKline Pharmaceuticals Ltd. |

\* preprint study

**Table 2:** Characterization of participants included in the studies, according to the evaluated alternative.

| Study | Compared alternatives | Number of participants | Mean age (SD). | Male n (%) | White n (%) | Comorbidities * n (%) | BMC $\geq 30$ kg/m<sup>2</sup> n (%) | Primary series of COVID-19 vaccine and/or boosters n (%) |
| --- | --- | --- | --- | --- | --- | --- | --- | --- |
| Ganatra et al., 2022 (11)<sup>a</sup> | Nirmatrelvir-ritonavir for 5 days. | 1,130 | 57.5 (16.3) | 418 (37.0) | 925 (81.9) | > 50% had at least 1 comorbidity. | 237 (21) | 1,130 (100) |
|  | Standard treatment | 1,130 | 57.7 (16.3) | 406 (35.9) | 941 (83.3) | > 50% had at least 1 comorbidity. | 208 (18) | 1,130 (100) |
| Yip et al., 2022 (12)<sup>a</sup> | Nirmatrelvir-ritonavir for 5 days. | 4,921 | 70.8 (12.1) | 2,247 (45.7) | NR | 1,970 (40) | 24.0 (4.2)<sup>b,d</sup> | 42.6 (15.8)<sup>e</sup> |
|  | No antiviral treatment. | 4,758 | 70.5 (12.2) | 2,178 (45.8) | NR | 1,907 (40) | 24.5 (4.7)<sup>b,d</sup> | 42.8 (15.7)<sup>e</sup> |
| Wai et al., 2023 (15) | Nirmatrelvir-ritonavir for 5 days. | 4,442 | 4,366 (98, | 2,016 (45.4) | NR<sup>f</sup> | > 10% had at least 1 comorbidity. | NR | NR |
|  | No antiviral treatment. | 23,430 | 21,904 (93.5%)<sup>c</sup> | 11,078 (47.3) | NR<sup>f</sup> | > 50% had at least 1 comorbidity. | NR | NR |
| Hedvat et al., 2022 (16) | Nirmatrelvir-ritonavir for 5 days. | 28 | 57.6 (44.3–68.6) | 11 (39.3) | NR | 28 (100) | 25.3 (22.3–30)<sup>b</sup> | 23 (82.1) |
|  | No antiviral treatment. | 75 | 53.3 (37.6–64.6) | 32 (42.7) | NR | 75 (100) | 27 (23.3–29.5)<sup>b</sup> | 61 (81.3) |
| Dryden-Peterson et al., 2022 (17)<sup>a</sup> | Nirmatrelvir-ritonavir for 5 days. | 11,797 | 50–64 years – 6,388 (54%)<br>≥ 65 years - 5,408 (46%) | 4,880 (41) | 10,164 (86) | ≤3: 6,727 (57)<br>≥4: 5,070 (43)<sup>i</sup> | 4,013 (34) | 10,752 (91) |
|  | No antiviral treatment. | 32,248 | 50–64 years - 17,881(55%)<br>≥ 65 years 14,367 (45%) | 12,603 (39) | 27,266 (85) | ≤3: 18,464 (57)<br>≥4: 13,784 (43)<sup>i</sup> | 10,661 (33) | 29,158 (90) |
| Wong et al., 2022 (18)<sup>a</sup> | Nirmatrelvir-ritonavir for 5 days. | 5,542 | 4,758 (85.9%)<sup>c</sup> | 2,566 (46.3) | NR | 0-4: 5291 (95.5)<sup>k</sup><br>5-14: 251 (4.5) | NR | 1,850 (33.4) |
|  | No antiviral treatment. | 54,672 | 46,601 (85.2%)<sup>c</sup> | 25,490 (46.6) | NR | 0-4: 52,345 (95.7)<br>5-14: 1,327 (4.3) | NR | 18,138 (33.2) |
| Arbel et. al, 2022 (19) | Nirmatrelvir-ritonavir for 5 days. | 3,902 | 67.4 (11.2) | 1,553 (40) | NR | 3,902 (100) | 1,626 (42) | 3,520 (90) |
|  | No antiviral treatment. | 105,352 | 59.6 (12.8) | 41,987 (40) | NR | 105,352 (100) | 36,140 (34) | 81,861 (78) |
| Schwartz et. al., 2023 (14)<sup>a</sup> | Nirmatrelvir-ritonavir for 5 days. | 8,876 | 74.3 (NI) | 3,613 (40.7) | NR | ≥ 3 3,805 (42.9)<br>< 3 5,071 (57.1) | NR | 8,326 (93.8) |
|  | No antiviral treatment. | 168,669 | 52.4 (NI) | 61,733 (36.6) | NR | ≥ 3 26,888 (15.9)<br>< 3 141,781 (84.1) | NR | 156,525 (92.8) |
| Aggarwal et al., 2023 (22)<sup>a</sup> | Nirmatrelvir-ritonavir for 5 days. | 7,168 | 18–44 years 3,288 (45.9%)<br>45–64 years 1,582 (22.1%)<br>≥ 65 years 2,298 (32.1%) | 2,966 (41.4) | 5,826 (81.3) | 4,378 (61.1) | 1,924 (26.8) | 5,416 (75.5) |
|  | No antiviral treatment. | 9,361 | 18–44 years 5,964 (63.7%)<br>45–64 years 1,442 (15.4%)<br>≥ 65 years 1,955 (20.9%) | 3,899 (41.7) | 7,365 (78.7) | 4,450 (47.5) | 1,793 (19.2) | 6,932 (74) |
| Najjar-Debbiny | Nirmatrelvir-ritonavir for | 4,737 | 68.5 (12.5) | 1,992 (42.1) | NR | 4,737 (100) | 1,938 (40.9) | 3,686 (77.8) |
| et al., 2022 (20) | 5 days. |  |  |  |  |  |  |  |
|  | No antiviral treatment. | 175,614 | 53.9 (16.8) | 71,967 (41.0) | NR | 175,614 (100) | 97,938 (55.8) | 131,796 (75.0) |
| Qian et al., 2022 (13) | Nirmatrelvir-ritonavir for 5 days. | 307 | 57.1 (14.9) | 72 (23.5) | 259 (84.4) | 260 (84.4) | 27.7 (7.3)<sup>b</sup> | 299 (97.4) |
|  | No antiviral treatment. | 278 | 58.3 (15.6) | 73 (26.3) | 223 (80.2) | 234 (84.2) | 27.0 (8.3)<sup>b</sup> | 260 (93.5) |
| Shah et al., 2022 (21) | Nirmatrelvir-ritonavir for 5 days. | 198,927 | 18–49 years 56,620 (28.5%)<br>50–64 years 66,929 (33.6%)<br>$\geq 65$ years 75,378 (37.9%) | 75,984 (38.2) | 158,696 (79.8) | 182,768 (91.9) | 98,892 (49.7) | 156,248 (78.5) |
| | No antiviral treatment. | 500,921 | 18–49 years 221,089 (44.1%)<br>50–64 years 147,274 (29.4%)<br>$\geq 65$ years 132,558 (26.5) | 184,184 (36.8) | 368,109 (73.5) | 463,849 (92.6) | 243,331 (48.6) | 325,058 (64.9) |
| Bajema et. al., 2022 (23)<sup>a</sup> | Nirmatrelvir-ritonavir for 5 days. | 1,587 | 65.0 (54.0,74.0) | 1,412 (89.0) | 1,111 (70.0) | 1,587 (100) | 818 (51.5) | 1,050 (66.3) |
|  | No antiviral treatment. | 1,587 | 66.0 (54.0,74.0) | 1,416 (89.3) | 1,149 (72.4) | 1,587 (100) | 817 (51.5) | 1,035 (65.2) |
| Lewnard et. al., 2023 (26) | Nirmatrelvir-ritonavir for 5 days. | 7,274 | 12–39 años -686 (9.4%)<br>40–59 años -2,659 (36.6%)<br>$\geq 60$ años 3,929 (54.0%) | 3,080 (42.3) | 1,921 (26.4) | 3,534 (48.6) | 3,253 (44.7) | 6,831 (93.9) |
| | No antiviral treatment. | 126,152 | 12–39 años -44,862 (35.6%)<br>40–59 años -49,864 (39.5%)<br>$\geq 60$ años 31,425 (24.9%) | 56,357 (44.7) | 26,884 (21.3) | 6,636 (21.1) | 39,482 (31.3) | 107,377 (85.1) |
| Zhou et. al., 2022 (27)<sup>a</sup> | Nirmatrelvir-ritonavir for 5 days. | 2,808 | 60.6 (15.8) | 1,183 (42.1) | 2,381 (84.8) | 1.38 (2.2)<sup>h</sup> | 1,214 (43.2) | 1,897 (67.6)<sup>g</sup> |
|  | No antiviral treatment. | 10,849 | 60.7 (16.7) | 4,539 (41.8) | 9,132 (84.2) | 1.36 (2.3)<sup>h</sup> | 4,870 (44.9) | 7,207 (66.4)<sup>g</sup> |
| Patel et al., 2022 (24) | Nirmatrelvir-ritonavir for 5 days. | 337 | 52.6 (15.5) | 178 (52.8) | 227 (67.4) | 337 (100) | 5 (1.5)<sup>j</sup> | 301 (89.3) |
|  | No antiviral treatment. | 4,044 | 52.4 (17.5) | 2,210 (54.7) | 1,986 (49.1) | 4,044 (100.0) | 72 (1.8)<sup>j</sup> | 3,488 (86.3) |

\* Cardiovascular diseases; digestive diseases; diabetes mellitus; malignant tumor; nervous system diseases; respiratory diseases; kidney diseases; HIV infection. SD: Standard Deviation; BMI: Body Mass Index; NR: Not Reported. a) Propensity Score Matching (PSM) or Weighted Analytic Cohort matched cohort; b) Mean BMI (SD); c) Studies by Wai et al., 2022, Wong et al., 2022 reported the number of patients over 60 and 65 years old (%); d) Yip et al, 2022, refers to BMI data before PSM; e) Rate of complete vaccination specified by age and sex (% and SD); f) 92.6% of the patients are of Chinese ethnicity.; g) Zhou et al., 2022, vaccination status was measured considering at least one dose ( $\geq 1$ dose); h) Deyo-Charlson Comorbidity Index Score; i) Dryden-Peterson et al., 2022 used the Monoclonal Antibody Screening Score - a comorbidity index that predicts the risk of hospitalization from COVID-19; j) Class 3 obesity: BMI $\geq 40$ kg/m<sup>2</sup>; k) Charlson Comorbidity Index score

### Study selection

After exporting a single Mendeley® file, the records were imported into Rayyan (8). Two independent researchers selected the records, and a third evaluator was consulted in case of doubts, both for screening (reading titles and abstracts) and eligibility (reading full texts).

The inclusion criteria for this systematic review were: (1) observational studies comparing the use of nirmatrelvir-ritonavir versus standard treatment or no antiviral treatment; (2) involving outpatients with COVID-19 at high risk of developing severe disease as the research population; and (3) determining death and/or hospitalization as the evaluated outcomes. No restrictions were imposed on publication date, language, or follow-up time. Studies reported only in conference proceedings were excluded.

The exclusion criteria were: (a) the study was a review article, letters to the editor, comments, consensus documents, clinical trials, pre-clinical studies, animal studies, or case reports; (b) the study did not focus on patients with COVID_19 or the diagnosis was unclear.

### Data extraction and quality assessment

Two independent researchers performed data extraction using a standardized collection method with Microsoft Office Excel®. A third review author fully checked all extracted data. The following information regarding the demographic characteristics of the studies was collected: first author, publication year, country, study design, general characteristics of the population, time of follow-up, predominant variant of SARS-CoV-2 at the time of the study, diagnostic criteria, number of participants per alternative compared, average age, proportion of male population, proportion of white population, comorbidities, body mass index (BMI), and COVID-19 vaccination status. Additionally, for dichotomous outcomes, data were collected on the number of patients with events in each compared alternative, odds ratio (OR), hazard ratio (HR), relative risk (RR), confidence interval (CI), or p-value.

The risk of bias was independently investigated by two researchers using the ROBINS-I tool, which assesses the risk of bias for non-randomized studies (9). Any discrepancies were resolved by consensus. To evaluate publication bias for the primary outcomes, visual inspection of the funnel plot was employed. The quality of evidence was assessed using the GRADE (Grading of Recommendations Assessment, Development and Evaluation) tool (10).

### Data synthesis and sensitivity analysis

The primary outcomes were hospitalization, mortality and the composite outcome of mortality and/or hospitalization within 35 days. Further subgroup analyses were conducted based on vaccination status and age group. To analyze the data, we used Review Manager® (RevMan) Version 5.4.1 (Review Manager, The Nordic Cochrane Centre, The Cochrane Collaboration, Copenhagen, Denmark). The heterogeneity of the results was assessed using the Cochran’s Q test and I^2^_statistic. If the p-value was less than .05 in the Q_statistic and I^2^_was ≥_50%, the heterogeneity was considered significant. We used the Mantel-Haenszel statistical method, the Sidik-Jonkman estimator for tau2, and the Hartung-Knapp adjustment for the random effects model to calculate pooled odd ratios (ORs) with corresponding 95% confidence intervals (CI). When numerical data were unavailable, we used the PlotDigitizer v3. 2022 free version to extract data from graphs. A sensitivity analysis was conducted to compare the published and unpublished studies, as well as those with and without techniques to adjust for patient characteristics (either through propensity score matching (PSM) or inverse probability treatment weighting (IPTW)).

To perform the meta-analyses, we assessed the homogeneity and transitivity by comparing the PICO abbreviations of each study (population inclusion and exclusion criteria, definitions of subpopulations, intervention and controls, and definitions of outcomes). As important discrepancies were identified, we discussed them as possible limitations of the meta-analyses.

We presented the characteristics of the studies, the characteristics of the participants, the individual results, and the methodological quality assessment of the included studies in a narrative and descriptive statistical form (absolute and relative frequency, mean and SD or median and interquartile range [IQR]), including tables to assist in the presentation of results. The narrative results were grouped by outcome, highlighting the alternatives compared.

## Results

### Search results and study selection

From the search strategy used, 182 publications were retrieved, with 162 citations remaining after identifying and eliminating duplicates. All records were subjected to a peer review process, and the full text of 32 potentially eligible articles was carefully considered. Of these 32 studies, 16 original articles were either not observational studies or did not have comparison groups. Therefore, records pertaining to sixteen (16) observational studies were included in the analysis. Fig 1 demonstrates the flow of our studies’ selection.

**Fig 1.**
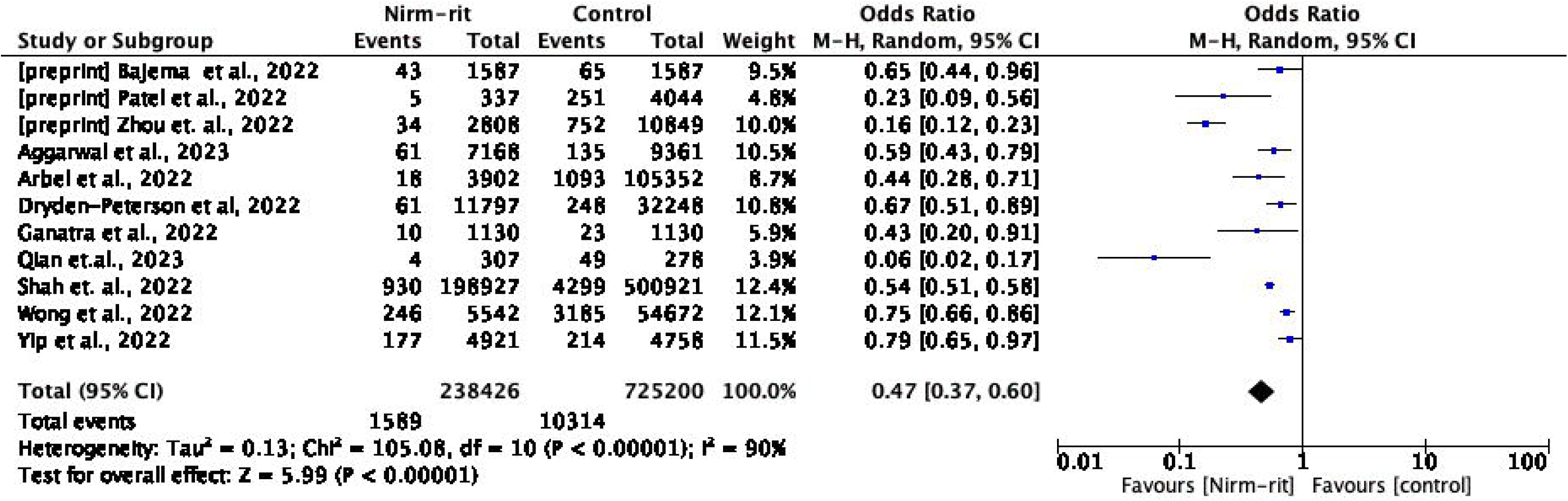
PRISMA Flow chart of literature screening.

### Study characteristics

The sixteen studies finally considered were conducted in 5 countries (Canada, China, United States, Israel, and United Kingdom). Of these, as of the last update of the search, 12 studies were published (11–22) and 4 were unpublished studies (preprint) (23–26). All studies were retrospective cohorts of data obtained from electronic records of hospitals and other healthcare centers, collected from January 2021 to October 2022.

For the meta-analysis, fourteen studies were considered. Data from the studies by Wai et al., 2022 (n=27,872) and Lewnard et al., 2023 (n=133,426) were not meta-analyzed, as the former did not contain all the necessary data for the proposed meta-analysis and the latter introduced a potential critical bias, as the evaluated cohort was a sample analysis where one or more baseline characteristics were retained in the evaluation, rather than all relevant baseline characteristics for an effectiveness assessment that make the groups minimally comparable. As a result, the cohort was still completely unbalanced (15,26).

All patients evaluated in the included studies, eligible for treatment with nirmatrelvir-ritonavir, met the high-risk criteria for progression to severe COVID-19 defined by their respective countries, which included criteria such as age, vaccination status, and presence of comorbidities. In the study by Aggarwal et al., 2023, the decision to seek antiviral treatment was made by patients and physicians, without necessarily meeting the eligibility criteria defined by the United States government (22).

Regarding the initiation of treatment with nirmatrelvir-ritonavir, 8 studies were strict with the initiation of treatment within the fifth day of symptom onset or positive COVID-19 test (11,12,17–20,24,25). In the other 6 studies, there was greater flexibility, as patients started treatment with nirmatrelvir-ritonavir within 10 days of symptom onset or positive test (15,16,21–23,26). This was not mentioned in the other two meta-analyzed studies.

In total, data from 1,482,923 patients from 14 studies were included in the meta-analysis. The characteristics of the included studies are shown in Tables 1 and 2.

### Risk of bias

The included studies were evaluated using the ROBINS-I tool, which assesses the risk of bias in non-randomized studies. The supporting information provides further details on the risk of bias assessments for studies that reported data on mortality, hospitalization, and the composite outcome of hospitalization or mortality. Regarding the mortality outcome, 4 of the 13 included studies had a low risk of bias, while 7 had a moderate risk. However, for the outcome of hospitalization within 35 days, 9 of the 11 studies were at risk of serious or critical bias, primarily due to outcome measurement bias (S1 file; Table 3). There was low risk of bias due to missing results or reporting bias.

**Table 3.** Effectiveness results of studies included in the review, by subgroups.

|  | < 60 years | ≥ 60 years | Primary series of COVID-19 vaccine and/or boosters n (%) | Non-vaccinated | Comorbidities | Without comorbidities |
| --- | --- | --- | --- | --- | --- | --- |
| **Hospitalization** |  |  |  |  |  |  |
| Aggarwal et al., 2023<sup>a</sup> | aOR: 0.53 (0.34-0.80) | aOR= 0.37 (0.23-0.57) | aOR=0.47 (0.29-0.74)<sup>d</sup> | aOR= 0.46 (0.27-0.77) | aOR: 0.37 (0.25-0.654) | aOR: 0.68 (0.41-1.12) |
| Arbel et al., 2022<sup>e</sup> | aHR: 0.74 (0.35 to 1.58) | aHR: 0.27 (0.15-0.49) | ≥ 65 años: aHR: 0.32 (0.17-0.63)<br>< 65 años: aHR: 1.13 (0.50-2.58) | ≥ 65 años: aHR: 0.15 (0.04-0.60)<br>< 65 años: aHR: 0.23 (0.03-1.67) | Not reported | NI |
| Shah et al., 2022 | 18-49: aHR: 0.59 (0.48-0.71)<br>50-64: aHR: 0.40 (0.34-0.58) | AHR: 0.53 (0.48-0.58) | ≥3 doses: aHR: 0.50 (0.45-0.55)<br>2 doses aHR: 0.50 (0.42-0.58) | aHR: 0.50 (0.43-0.59) | 1 aHR: 0.57 (0.45-0.71)<br>≥2 aHR: 0.47 (0.44-0.51) | aHR: 0.89 (0.58-1.36) |
| Qian et al., 2022 | aOR: 0.07 (0.02-0.31) | aOR: 0.11 (0.02-0.54) | aOR: 0.09 (0.03 - 0.32) | Not reported | Not reported | Not reported |
| Yip et. al., 2022 | Not reported | aHR: 0.76 (0.63-0.92)<sup>a</sup> | Not reported | Not reported | aHR: 0.76 (0.63-0.92)<sup>c</sup> | Not reported |
| Zhou et al., 2022 | aHR: 0.19 (0.09, 0.38) | aHR: 0.17 (0.12, 0.26) | aHR: 0.18 (0.12, 0.28) | NI | Not reported | Not reported |
| Wong et al., 2022 | HR: 0.50 (0.31, 0.81) | HR: 0.80 (0.69, 0.91) | HR: 0.71 (0.51, 1.01) | HR: 0.76 (0.66, 0.87) | Not reported | Not reported |
| Ganatra et al., 2022 | Not reported | Not reported | 0.43 (0.2-0.9) | Not reported | Not reported | Not reported |
| **Mortality** |  |  |  |  |  |  |
| Schwartz et al., 2022<sup>d</sup> | OR: 0.13 (0.03 - 0.57)<sup>f</sup> | OR: 0.48 (0.39 - 0.59) | 1-2 doses: OR 0.23 (0.11 - 0.51)<br>3+ doses: OR 0.54 (0.43 - 0.67) | OR 0.34 (0.16 - 0.74) | 3+: 0.48 (0.34 - 0.67)<br><3: 0.50 (0.39 - 0.64) | Not reported |
| Arbel et al., 2022<sup>b</sup> | aHR: 1.32 (0.16-10.75) | aHR: 0.21 (0.05-0.82) | Not reported | Not reported | Not reported | Not reported |
| Wong et al., 2022 | Not reported | HR: 0.48 (0.32, 0.74) | Not reported | HR: 0.44 (0.30, 0.66) | Not reported | Not reported |
| **Mortality or hospitalization** |  |  |  |  |  |  |
| Najjar-Debbiny et al., 2022 | aHR: 1.06 (0.36-0.73) | aHR: 0.52 (0.36-3.15) | aOR: 0.62 (0.39-0.98) | aOR: 0.52 (0.32-0.82) | Not reported | Not reported |
| Dryden-Peterson et al, 2022 | aRR: 0.55 (0.30 - 1.03) | aRR: 0.55 (0.40 to 0.77) | aRR: 0.69 (0.50 - 0.94) | aRR: 0.19 (0.08 - 0.49) | aRR: 0.56 (0.40 to 0.78)<sup>e</sup> | Not reported |
| Bajema et al., 2022 | aRR: 0.81 (0.46-1.42) | aRR: 0.46 (0.31-0.66) | aRR: 0.48 (0.32-0.73) | aRR:0.61 (0.38-0.97) | Not reported | Not reported |
| Lewnard et. al., 2022 | Not reported | Not reported | HR: 0.45 (0.21 - 0.94) | Not reported | Not reported | Not reported |
| Schwartz et al., 2023<sup>f</sup> | OR 0.34 (0.15 - 0.79) | OR 0.55 (0.45 - 0.66) | 1-2 doses: OR 0.25 (0.12 - 0.50)<br>3+ doses: OR 0.62 (0.51 - 0.75) | OR 0.44 (0.23 - 0.84) | 3+: 0.54 (0.39 - 0.73)<br><3: 0.57 (0.46 - 0.71) | Not reported |

a) Aggarwal et al., 2023 – considered comorbidities 0-1 as with comorbidities; b) Arbel et al., 2022 defined previous immunity to SARS-CoV-2 as previous vaccination or SARS-CoV-2 infection while the rest of the studies defined it only as previous vaccination; c) Yip et al. al evaluated >60 years or <60 years with comorbidity; d) Schwartz et al., 2022 analyzed age groups < and > 70 years; e) Dryden-Peterson et al, 2022, Monoclonal Antibody Screening Score ≥ 4; f) iSchwartz et al., 2022 analyzed age groups of < and > 70 years

### Effectiveness outcomes

Table 3 shows the effect measures reported by the studies included in this review, stratified by subgroup. In the supplementary material (S1 file, Table 4), we report the aggregated results reported and used in the meta-analysis. The following are the results of the meta-analyses conducted by the evaluated outcome.

**Table 4.**
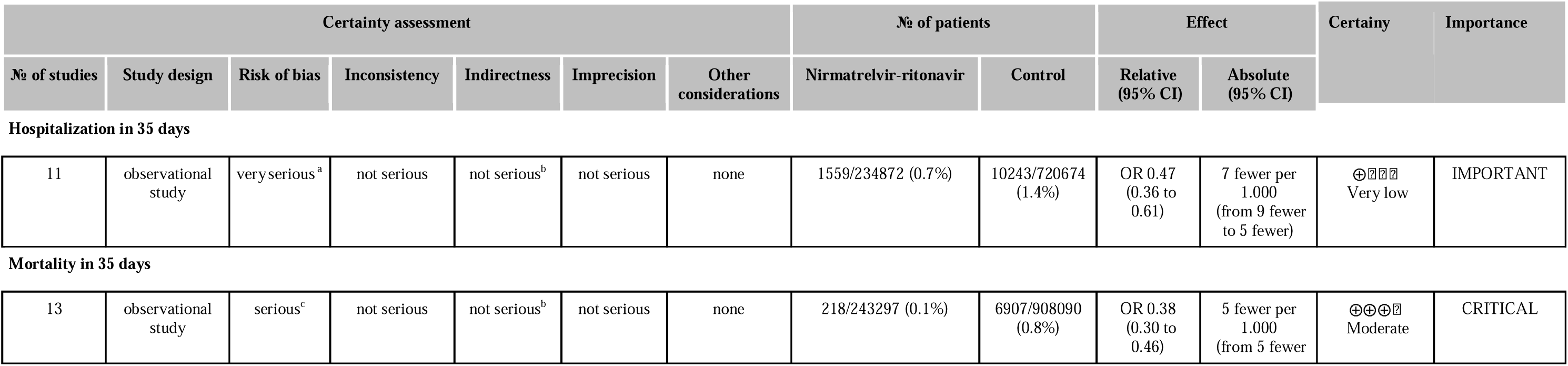
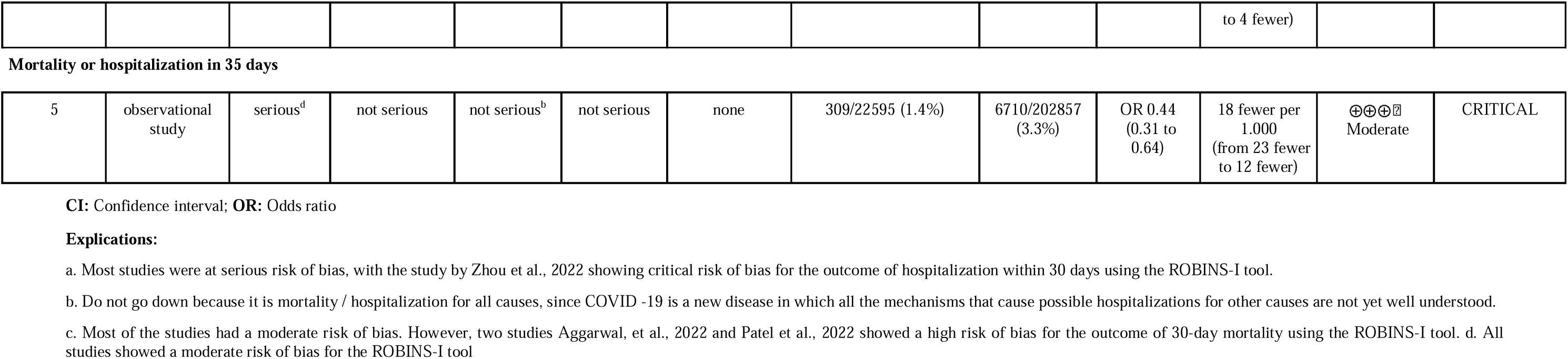
Summary of evidence about treatment with nirmatrelvir-ritonavir versus standard treatment (without antivirals) for COVID-.

#### Mortality

Thirteen studies reported mortality data, including 1,159,467 patients and 7,133 deaths (11,13–19,21–25). In comparison to standard treatment without antivirals, nirmatrelvir-ritonavir reduced the risk of death by 62% (OR= 0.38; 95% CI: 0.30-0.46; moderate certainty of evidence) (Fig 2).

**Fig 2.**
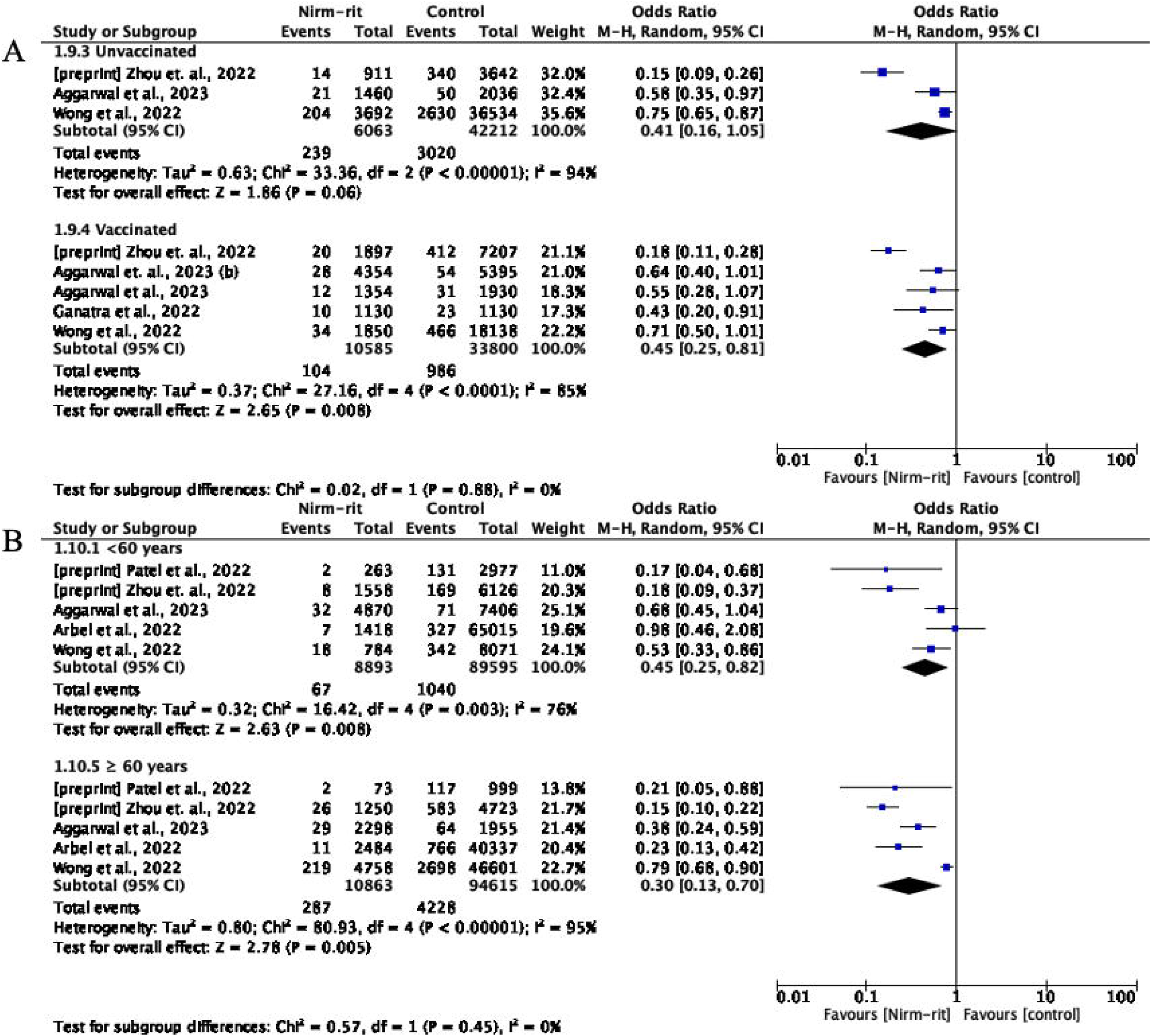
Forest plot of all-cause mortality outcome within 35 days - nirmatrelvir-ritonavir versus control.

Three studies reported subgroup data by vaccination status (11,14,18) and four other studies reported data by age group (14,18,19,24). In the analysis by vaccination status, nirmatrelvir-ritonavir reduced the risk of mortality both in the unvaccinated group (OR=0.41; 95% CI: 0.29-0.58) and in the vaccinated group (OR=0.31; 95% CI: 0.14-0.68), with no significant difference between the groups (Fig 3A).

**Fig 3.**
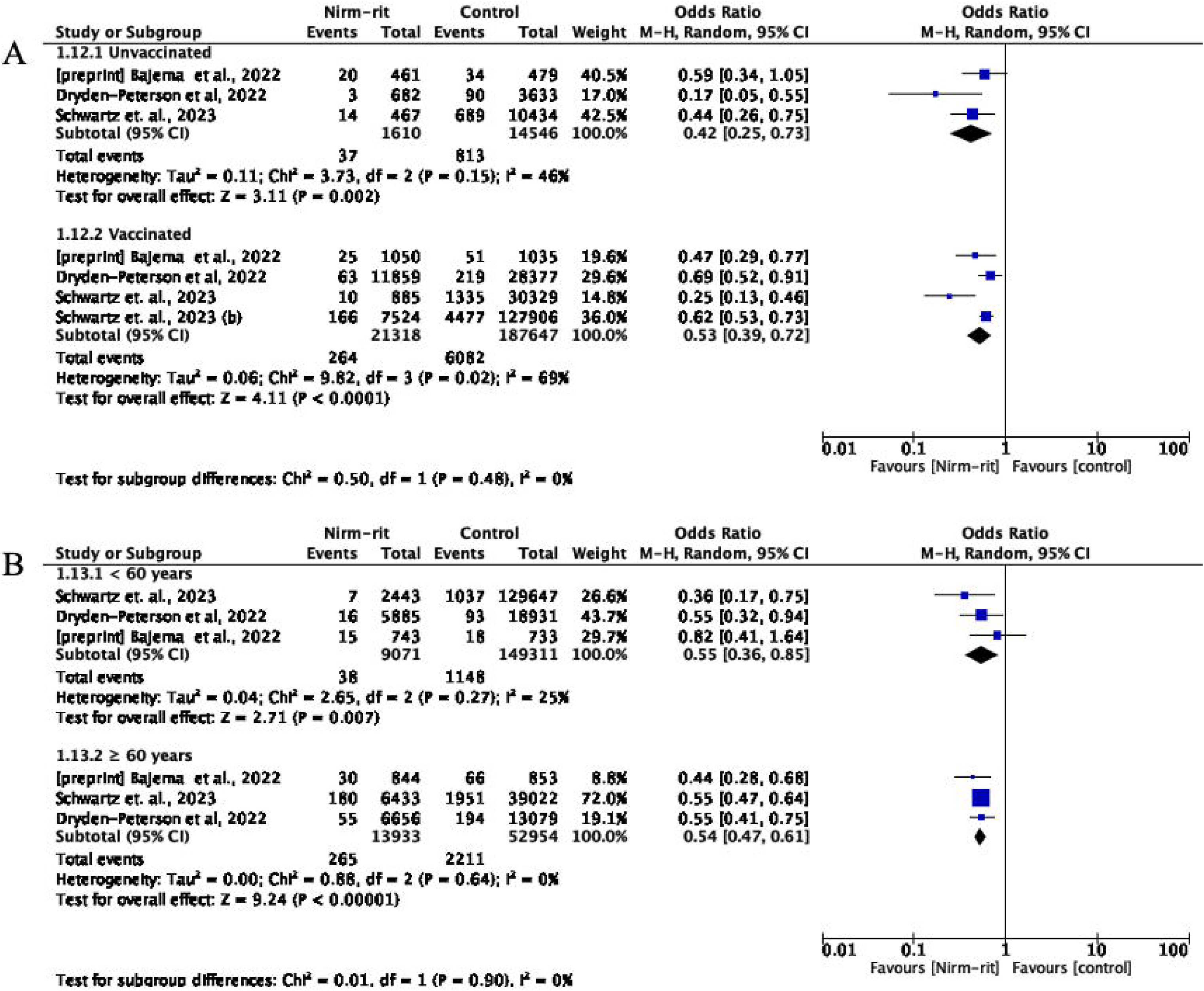
A: Forest plot of all-cause mortality outcome by vaccination status subgroup. B: Forest plot of all-cause mortality outcome by subgroup of age group.

In the subgroup of patients under 60 years of age, there appears to be no difference between treatment with nirmatrelvir-ritonavir compared to standard treatment (OR=0.48; 95% CI: 0.09-2.50), while treating patients over 60 years of age with nirmatrelvir-ritonavir suggests greater protection against the risk of death (OR=0.47; 95% CI: 0.40-0.55) (Fig 3B).

It should be noted that the subgroup meta-analysis could only be performed among those studies that reported data that could be grouped. Table 3 presents the results of the effect measures from other studies that reported the evaluation of these subgroups.

Furthermore, the sensitivity analysis did not reveal significant changes in the mortality rate of published studies (OR = 0.42; 95% CI: 0.35–0.50) and unpublished studies (OR = 0.23; 95% CI: 0.13–0.42). There were also no significant differences between matched studies (OR = 0.34; 95% CI: 0.25–0.47) and unmatched studies (OR = 0.38; 95% CI: 0.27–0.54) (S1 file: Fig 1 and Fig 2).

#### Hospitalization

Eleven studies reported data on hospitalization within 35 days of follow-up after the initiation of the treatment, which included 963,626 patients, with the occurrence of 11,903 events (11–13,17–19,21–25)

Compared to standard treatment or no antiviral treatment, the use of nirmatrelvir-ritonavir resulted in a 53% reduction in the risk of hospital admission (OR = 0.47; 95% CI: 0.37–0.60, with very low certainty of evidence) (Fig 4).

**Fig 4.**
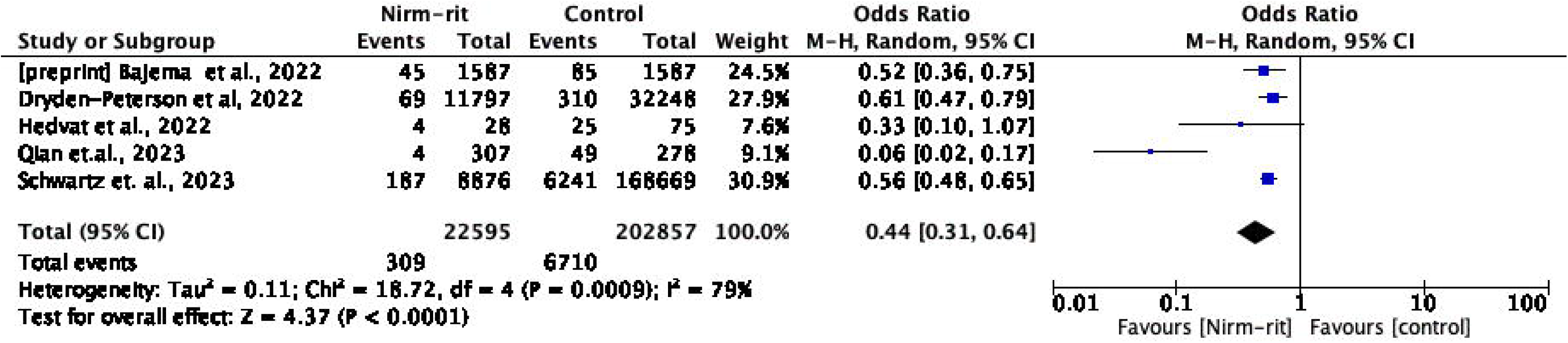
Forest plot of all-cause hospitalization outcome within 35 days - nirmatrelvir-ritonavir versus control.

Four studies reported subgroups data by vaccination status (11,18,22,25) and five studies reported age subgroups data (18,19,22,24,25). In the subgroup analysis of state vaccination, nirmatrelvir-ritonavir reduced the risk of hospitalization in both groups, non-vaccinated (OR= 0.41; 95%CI: 0.16-1.05) and vaccinated (OR=0.45; 95%CI: 0.25-0.81). It is worth noting that when using the random effects method, the meta-analysis result introduced greater inaccuracy in the data. Although each study showed a reduction in risk favoring the treatment of nirmatrelvir-ritonavir in the non-vaccinated group, the effect magnitude was very different between the studies in this analysis. In the subgroup analysis by age, nirmatrelvir-ritonavir reduced the risk of hospitalization in both the group of individuals under 60 years (OR=0.45; 95%CI: 0.25–0.82) and the group of individuals over 60 years (OR=0.30; CI95%: 0.13–0.70), without a significant difference between the two groups (Fig 5).

**Fig 5.**
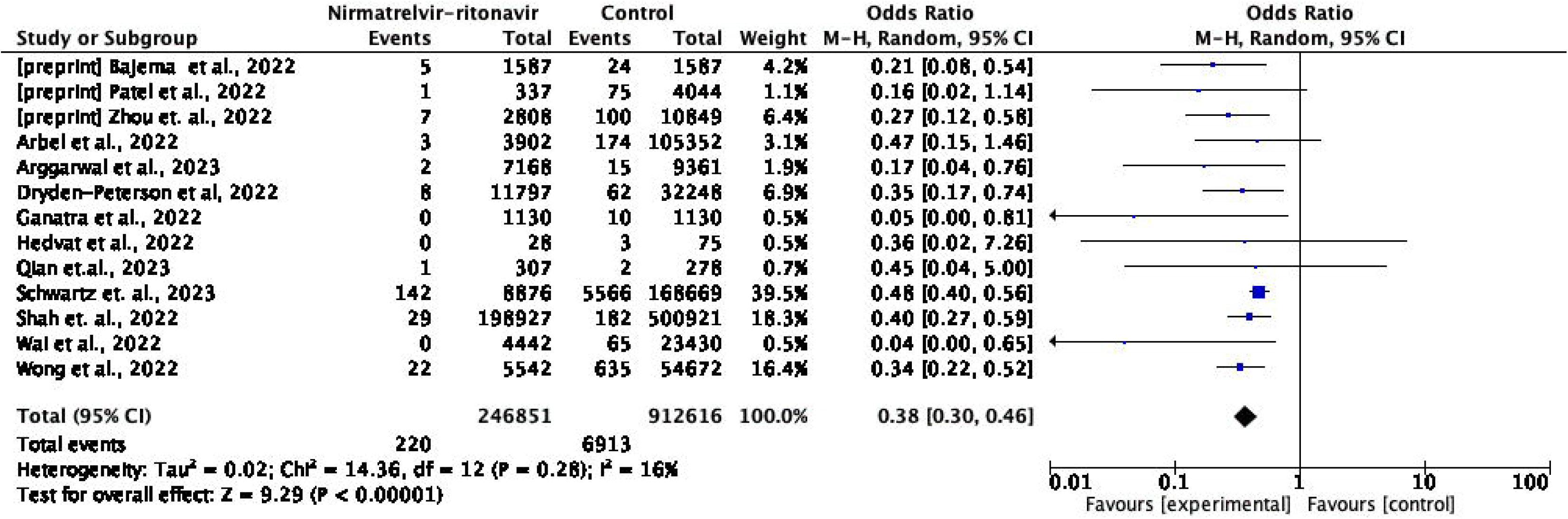
A: Forest plot of all-cause hospitalization outcome within 35 days by vaccination status subgroup. B: Forest plot of all-cause hospitalization outcome within 35 days by subgroup of age group.

The sensitivity analysis revealed significant changes in the hospitalization rate between published studies (OR = 0.57; 95%CI: 0.46–0.71) and unpublished studies (OR = 0.29; 95 %CI: 0.10–0.84). There were also differences between adjusted studies (OR = 0.52; 95 %CI: 0.37–0.73) and not adjusted (OR = 0.29; 95 %CI: 0.15–0.56) (S1 file: Fig 3 and Fig 4).

#### Outcome composed of mortality and/or hospitalization

Five studies reported effectiveness data based on the outcome composed of mortality and/ or hospitalization within 35 days of follow-up after the start of treatment, which included 225,452 patients, with the occurrence of 7,019 events (13,14,16,17,23)

Compared to standard treatment or no antiviral treatment, nirmatrelvir-ritonavir reduced the risk of mortality or hospitalization by 56% (OR = 0.44; 95% IC: 0.31–0.64, moderate certainty of evidence) (Fig 6).

**Fig 6.**
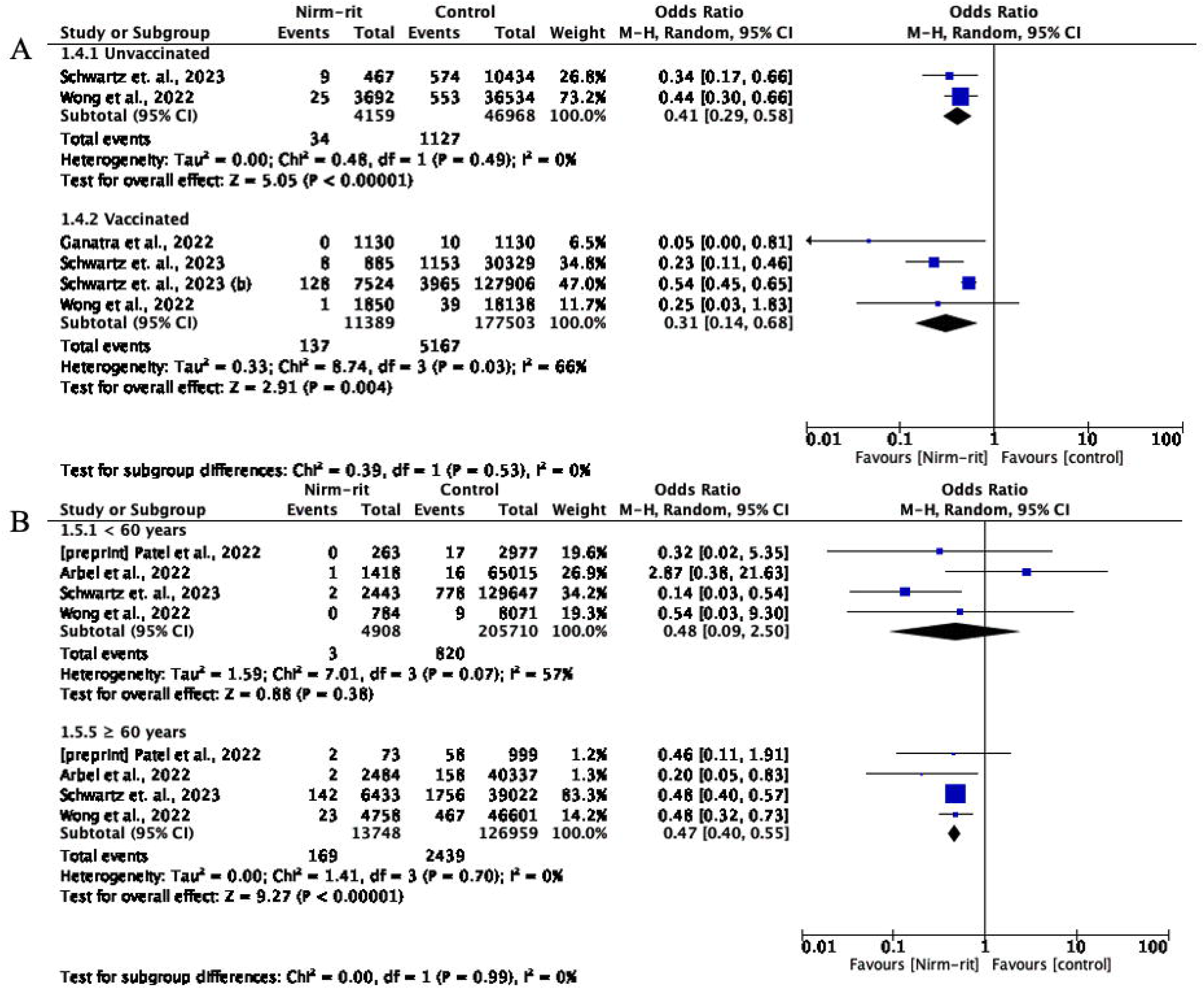
Forest plot of all-cause mortality or hospitalization outcome within 35 days – nirmatrelvir-ritonavir versus control.

In the subgroup analysis of vaccinated and non-vaccinated individuals, the treatment with nirmatrelvir-ritonavir reduced the risk of mortality or hospitalization by 47% (OR= 0.53; 95%CI: 0.39–0.72) and 58% (OR= 0.42; 95%CI: 0.24–0.73), respectively.

Among patients under 60 years of age, nirmatrelvir-ritonavir reduced the risk of mortality or hospitalization by 45% (OR= 0.55; 95%CI: 0.36–0.85), while in patients over 60 years of age, it reduced the risk by 46% (OR= 0.54; 95%CI: 0.47–0.61) (Fig 7).

**Fig 7.**
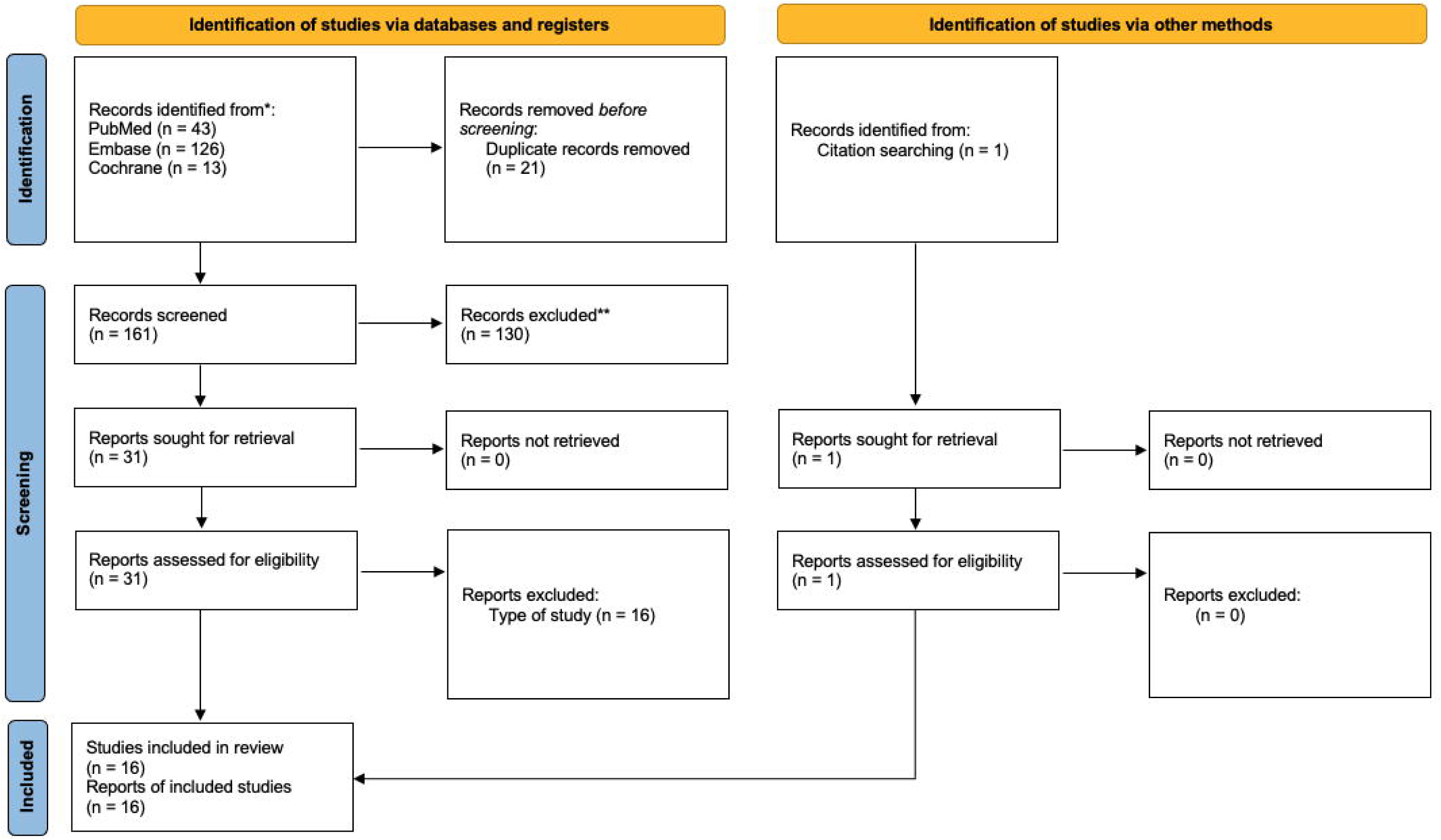
A: Forest plot of hospitalization or mortality outcome within 35 days by vaccination status subgroup. B: Forest plot of hospitalization or mortality by subgroup of age group.

### Certainty of the evidence

The GRADE tool (Grading of Recommendations Assessment, Development and Evaluation) was utilized to assess the quality of evidence. A total of 16 studies were included as evidence, with 14 of these being meta-analyzed for the three primary outcomes of interest. All studies demonstrated significant results in reducing the risk of death and/or hospitalization with the use of nirmatrelvir-ritonavir (Table 4).

Regarding the hospitalization outcome within 35 days, the majority of studies exhibited a high risk of bias, thus the overall bias risk domain was considered very serious. The domain of inconsistency was also rated as serious, despite the absence of contrasting results, as the summary of study results revealed considerable heterogeneity (I^2^ = 92%, p < 0.00001). Conversely, the remaining domains were classified as non-serious due to the absence of studies with discrepant results, and we consider that the summary result was not subject to significant imprecision.

In relation to mortality outcomes within 35 days and mortality or hospitalization within 35 days, the majority of studies exhibited a moderate risk of bias and therefore the global risk of bias domain was considered serious. However, the remaining domains were considered non-serious, due to the absence of discrepant results and we considered that the summary result had no important imprecision.

Moreover, despite acknowledging that most studies measured mortality and hospitalization outcomes for all causes rather than specifically for COVID-19, it was determined that the domain of indirect evidence would be classified as non-serious for all outcomes. This decision was made due to COVID-19 being a novel disease with poorly elucidated mechanisms, which means that certain hospitalizations and deaths for all causes may be directly linked to COVID-19.

Regarding factors that can increase the quality of the evidence, we assessed the publication bias of the main outcome measures by qualitatively evaluating the funnel plot. No significant asymmetries were detected, leading us to conclude that there was no suspicion of publication bias. Since all the included studies used the same dose of nirmatrelvir-ritonavir, it was not possible to detect a dose-response gradient. We considered that there was no residual confounding effect from observational studies that could reduce or increase the demonstrated effect. Moreover, we determined that the magnitude of the effect was not sufficiently large to increase the quality of the evidence.

## Discussion

The aim of this systematic review and meta-analysis was to evaluate the effectiveness of nirmatrelvir-ritonavir treatment in real-world situations, using observational studies that considered different scenarios of the target population, who were at high risk of hospitalization, such as vaccination status, age group, presence of comorbidities, and other associated risk factors in patients with mild to moderate COVID-19.

This study found that nirmatrelvir-ritonavir treatment was linked to a decreased risk of hospitalization and mortality, which is consistent with the results of previous reviews conducted by Amani B et al. and Cheema et al. (28,29). In the same direction as these results, although with a different magnitude, Hammond et al. conducted a phase 2-3 clinical trial (EPIC-HR) to evaluate the efficacy and safety of nirmatrelvir-ritonavir for non-hospitalized adult patients with mild to moderate COVID-19 at high risk of severe illness, resulting in an 88.9% relative risk reduction of hospitalization or death (30). The differences observed in the effectiveness of nirmatrelvir-ritonavir treatment across different populations and contexts reflect the challenges posed by significant interindividual variations in COVID-19. These variations can be influenced by factors such as individual risk, the several mutations in coronavirus genotypes (variants), vaccination coverage, geographic location, and healthcare systems, and can impact hospitalization criteria, timing, and treatment effectiveness. In addition to inherent variations in study methodology, these factors make it challenging to compare studies results across different populations and contexts (31–34). This also means that the issue of discrepancies between results from randomized controlled trials (RCTs) and observational studies can be explained by the obvious efficacy-effectiveness gap and should not promote direct comparisons (35).

Aligned with the main findings, subgroup analyses comparing vaccinated and unvaccinated patients indicated a significant reduction in the risk of mortality and hospitalization. Despite the varied vaccination status of the studies included in this review, it was observed that some high-risk patients did not receive a COVID-19 vaccine. In this group, treatment with nirmatrelvir-ritonavir may confer protection against mortality and hospitalization. It is also important to consider that despite the immunological escape of the Omicron variant, the vaccines still provide important protection against COVID-19 (36,37). Moreover, the Omicron variant of COVID-19 has been demonstrated to have lower rates of hospitalization and mortality compared to previous variants. These factors can affect the effect of treatment with Nirmatrelvir-ritonavir (38,39). Additionally, the efficacy of nirmatrelvir-ritonavir use within the context of the availability of bivalent COVID-19 vaccines requires further consideration and evaluation.

Our meta-analysis results by age group indicate that nirmatrelvir-ritonavir treatment may provide benefits for both younger and older COVID-19 patients in terms of hospitalization and composite outcome of mortality or hospitalization, suggesting that the findings of this study may be applicable to a broad population. However, in terms of mortality for population under 60 years, the risk reduction could not be confirmed by the meta-analysis. A separate study conducted by Arbel et al., found that only high-risk COVID-19 positive outpatients aged 65 years and older experienced reduced deaths and hospitalizations with nirmatrelvir-ritonavir treatment. The possible reasons that explain this difference include the study period, taking into account the new variants of COVID-19, hospitalization criteria for young patients, vaccination status, and presence of comorbidities (19,22). This review suggest that nirmatrelvir-ritonavir is effective in treating non vaccinated or vaccinated, non-severe COVID-19 patients with high risk for hospitalization. This may have potential implications for clinicians and decision-makers and could alleviate the pressure on the healthcare system due to COVID-19 hospitalizations. The living clinical guideline developed by the WHO makes a strong recommendation in favor of nirmatrelvir-ritonavir as the first-choice treatment for non-severe patients with a high risk of hospital admission, and the recent update recommends treatment for pregnant and lactating women as well (4). Another COVID-19 antiviral, molnupiravir (Lagevrio®) got a refusal of the marketing authorization by the European Medicines Agency (EMA) on the grounds that the risk-benefit balance could not be established and that it was not possible to identify a specific group of patients in which a clinically relevant benefit could be demonstrated (40). In this scenario, the therapeutic arsenal for treating COVID-19 is more restricted.

Treating non-severe patients might be of interest, considering that antiviral drugs may be more useful in non-severe cases of COVID-19, where viral replication is the primary mechanism driving disease progression. This contrasts with severe cases, where the primary cause of illness is an inflammatory response (41–43). Furthermore, a randomized clinical trial conducted by Liu et al. in 2023, which evaluated the efficacy of nirmatrelvir-ritonavir in adult patients hospitalized with SARS-Cov-2 (Omicron BA.2.2 variant) infection and severe comorbidities, did not show any additional benefits in terms of all-cause mortality up to day 28 when compared to standard treatment (39).

The strengths of our systematic review are several. Firstly, only ambulatory patients considered at high risk of hospitalization were included in the review. Secondly, we conducted subgroup analyses by vaccination status and age group. Thirdly, we updated the data from the included preprint studies that had been published at the time of article writing. Additionally, the study was conducted in accordance with PRISMA guidelines, with the assessment of the risk of bias according to ROBINS-I and the GRADE assessment of available evidence. We conducted our search accounting for the latest publications with broad geographical distribution. To our knowledge, this is the first systematic review with meta-analysis that highlights differences in vaccination status, age group, and comorbidity presence. Our review included studies with heterogeneous populations as compared to the EPIC-HR trial, where 71% of the participants were Caucasians and the high-risk patients were mostly obese. This heterogeneity increases the external validity of our results.

Our systematic review also has some limitations. Firstly, all the studies included were retrospective cohorts, which are more susceptible to bias and confounding. However, to mitigate this limitation, most of the studies were matched by propensity score or other balancing methods between groups. Additionally, all the studies underwent assessment by the ROBINS-I bias risk tool, which enabled us to conduct a more rigorous evaluation and determine the confidence of the results using the GRADE method (10). Despite these efforts, the high heterogeneity between the studies and the subgroups evaluated, especially for the outcome of hospitalization within 35 days, suggests the possibility of variations in criteria for patient hospitalization decisions, different COVID-19 variants, patient characteristics, geographical location, and other factors (33,34).

A further limitation is that standard treatment or no use of antiviral treatment was considered as the control group in the studies. This may have affected the reported effect size and should be considered when interpreting our results (4).

Another limitation of our study is that only a few studies could be meta-analyzed by subgroup, which may distort the actual effect in these specific groups. To address this limitation, we reported effect measures adjusted by studies that conducted such analyses but were not included in the meta-analysis due to the absence of data.

The timing of antiviral therapy initiation is a critical consideration for the management of COVID-19 patients. The World Health Organization recommends starting treatment within five days of symptom onset (4). However, in the studies we analyzed, the duration of symptoms or the date of positive COVID-19 test before treatment initiation varied widely (up to 10 days), and data on the timing of treatment initiation was often unavailable in some studies. This lack of data poses challenges in interpreting our findings regarding the optimal timing of oral antiviral therapy initiation. Nevertheless, the available evidence suggests that delaying the initiation of nirmatrelvir-ritonavir therapy beyond five days of symptom onset significantly reduces treatment efficacy against hospitalization and death (26,44). It is important to highlight that the beginning of treatment should be accompanied by early diagnosis, and therefore, it is crucial that countries have access to and implement efficient testing programs, especially in low- and middle-income countries (45).

Safety data, rebound effect and long-term outcomes of COVID-19 reported in some studies were not included in our analysis. Hammond et al, demonstrated a lower frequency of serious adverse events, and adverse events leading to discontinuation in the Nirmatrelvir-ritonavir group compared to the placebo group. Similarly, the systematic review by Amani et al., demonstrated that there was no significant difference in the incidence of adverse events between the treatment and control groups in their pooled analysis (OR = 2.20; 95% CI: 0.42–11.47) (28,30). In addition, it should be noted that ritonavir is a CYP3A4 inhibitor, an enzyme responsible for metabolizing several medications, and potential drug interactions should be taken into consideration during treatment, especially among poly-treated patients and those who are taking corticosteroids and other immunosuppressive medications (46).

Retrospective studies have suggested a low incidence of rebound phenomenon after treatment with nirmatrelvir-ritonavir, which was described in a limited number of individuals, all of whom developed virological rebound approximately between 7 and 30 days after symptom onset and were likely infected with Omicron variants. Among patients who developed symptom rebound after treatment with nirmatrelvir-ritonavir, the clinical presentation was mild and did not require COVID-19 directed therapies (28,47–50). It should be noted that prospective epidemiological studies are still needed to more accurately measure the incidence and risk factors for COVID-19 rebound and compare them in those treated with nirmatrelvir-ritonavir versus those not treated.

Finally, considering the potential benefits of treatment with nirmatrelvir-ritonavir and the necessary precautions to guide treatment. There are challenges to consider in the healthcare systems of countries, given that it is an expensive treatment with limited availability. There is a need to further evaluate prioritization, cost-effectiveness and the impact of its use, especially in low and middle-income countries (51,52).

## Conclusion

The results of our meta-analysis suggest that nirmatrelvir-ritonavir could be effective in reducing hospitalization and/or mortality in high-risk individuals with COVID-19, compared to those who did not receive antiviral treatment, either vaccinated or unvaccinated. Although it is important to mention that the effect on mortality reduction was uncertain for those under 60 years. The present review underscores the critical importance of early initiation of antiviral therapy. It is crucial to acknowledge that there are still several limitations to consider, and additional evidence is necessary to identify the subgroups of patients who may benefit the most from this treatment. It is important to highlight that observational studies are more prone to bias and confounding, and therefore cannot provide conclusive evidence of causality. Data from ongoing and future randomized controlled trials may further expand our understanding of the efficacy and safety of nirmatrelvir-ritonavir and help improve standard treatment guidelines for COVID-19.

## Data Availability

All relevant data are within the manuscript and its Supporting Information files.

## Supporting information

**S1 File – Contains PRISMA checklist, supporting materials, tables and figures**

