## Supplementary material for "Effectiveness of Nirmatrelvir-Ritonavir for the treatment of patients with mild to moderate COVID-19 and at high risk of hospitalization: Systematic review and meta-analyses of observational studies": S1 File

### Supplemental Table 1 – Search strategy

**PubMed:**

(((((("nirmatrelvir and ritonavir drug combination"[Supplementary Concept]) OR ("PF-07321332, ritonavir"[Title/Abstract])) OR ("nirmatrelvir, ritonavir"[Title/Abstract])) OR (Paxlovid[Title/Abstract])) OR (PF-07321332, ritonavir[Title/Abstract])) OR (nirmatrelvir, ritonavir drug combination[Title/Abstract])) AND (("Observational Studies as Topic"[Mesh] OR "Observational Study" [Publication Type] OR "Cohort Studies"[Mesh] OR "Longitudinal Studies"[Mesh] OR "Follow-up Studies"[Mesh] OR "Prospective Studies"[Mesh] OR "Retrospective Studies"[Mesh] OR "Case-Control Studies"[Mesh] OR "Cross-Sectional Studies"[Mesh] OR "Follow-up studies"[Mesh]) OR ("Observational Study"[Title/Abstract] OR "Observational Studies"[Title/Abstract] OR Cohort*[Title/Abstract] OR "Longitudinal study"[Title/Abstract] OR "Longitudinal Studies"[Title/Abstract] OR "Retrospective study"[Title/Abstract] OR "Retrospective studies"[Title/Abstract] OR "Prospective study"[Title/Abstract] OR "Prospective studies"[Title/Abstract] OR "Follow-up study"[Title/Abstract] OR "Follow-up studies"[Title/Abstract] OR "Case-Control Study"[Title/Abstract] OR "Case-Control Studies"[Title/Abstract] OR "Case-Comparison Studies" [Title/Abstract] OR "Case-Comparison Study"[Title/Abstract] OR Cross-sectional[Title/Abstract] OR "Transversal study" [Title/Abstract]))

**EMBASE:**

('coronavirus disease 2019'/exp OR 'coronavirus disease 2019') AND ('nirmatrelvir plus ritonavir' OR paxlovid OR 'nirmatrelvir') AND ('observational studies as topic' OR 'cohort studies' OR 'cross-sectional studies' OR 'observational study' OR 'observational studies' OR cohort OR 'longitudinal study' OR 'longitudinal studies' OR 'retrospective study' OR 'retrospective studies' OR 'prospective study' OR 'prospective studies' OR 'follow-up study' OR 'follow-up studies' OR 'case-control study' OR 'case-control studies' OR 'case-comparison studies' OR 'case-comparison study' OR 'cross sectional' OR 'transversal study')

**The Cochrane Library (reviews only):**

| | |
|---|---|
| #1 | MeSH descriptor: [COVID-19 ] explode all trees |
| #2 | "PF-07321332, ritonavir" |
| #3 | "nirmatrelvir, ritonavir" |
| #4 | Paxlovid |
| #5 | nirmatrelvir |
| #6 | "PF 07321332" |
| #7 | #2 OR #3 OR #4 OR #5 OR #6 |
| #8 | #1 AND #7 |

### Supplemental Table 2 – PRISMA checklist

| Section and Topic | Item # | Checklist item | Location where item is reported |
|---|---|---|---|
| **TITLE** | | | |
| Title | 1 | Effectiveness of Nirmatrelvir-Ritonavir for the treatment of patients with mild to moderate COVID-19 and at high risk of hospitalization: Systematic reviews and meta-analyses of observational studies | Page 1 |
| **ABSTRACT** | | | |
| Abstract | 2 | See the PRISMA 2020 for Abstracts checklist. | Page 2 |
| **INTRODUCTION** | | | |
| Rationale | 3 | Describe the rationale for the review in the context of existing knowledge. | Page 3-5 |
| Objectives | 4 | Provide an explicit statement of the objective(s) or question(s) the review addresses. | Page 6 |
| **METHODS** | | | |
| Eligibility criteria | 5 | Specify the inclusion and exclusion criteria for the review and how studies were grouped for the syntheses. | Page 6-7 |
| Information sources | 6 | Specify all databases, registers, websites, organizations, reference lists and other sources searched or consulted to identify studies. Specify the date when each source was last searched or consulted. | Page 6 |
| Search strategy | 7 | Present the full search strategies for all databases, registers and websites, including any filters and limits used. | Supplementary Material |
| Selection process | 8 | Specify the methods used to decide whether a study met the inclusion criteria of the review, including how many reviewers screened each record and each report retrieved, whether they worked independently, and if applicable, details of automation tools used in the process. | Page 6-7 |
| Data collection process | 9 | Specify the methods used to collect data from reports, including how many reviewers collected data from each report, whether they worked independently, any processes for obtaining or confirming data from study investigators, and if applicable, details of automation tools used in the process. | Page 6-8 |
| Data items | 10a | List and define all outcomes for which data were sought. Specify whether all results that were compatible with each outcome domain in each study were sought (e.g. for all measures, time points, analyses), and if not, the methods used to decide which results to collect. | Page 6-8 |
| | 10b | List and define all other variables for which data were sought (e.g. participant and intervention characteristics, funding sources). Describe any assumptions made about any missing or unclear information. | Page 7-8 |
| Study risk of bias assessment | 11 | Specify the methods used to assess risk of bias in the included studies, including details of the tool(s) used, how many reviewers assessed each study and whether they worked independently, and if applicable, details of automation tools used in the process. | Page 7 |
| Effect measures | 12 | Specify for each outcome the effect measure(s) (e.g. risk ratio, mean difference) used in the synthesis or presentation of results. | Page 7-8 |
| Synthesis methods | 13a | Describe the processes used to decide which studies were eligible for each synthesis (e.g. tabulating the study intervention characteristics and comparing against the planned groups for each synthesis (item #5)). | Page 7-8 |
| | 13b | Describe any methods required to prepare the data for presentation or synthesis, such as handling of missing summary statistics, or data conversions. | Page 7 |
| | 13c | Describe any methods used to tabulate or visually display results of individual studies and syntheses. | Page 7 |
| | 13d | Describe any methods used to synthesize results and provide a rationale for the choice(s). If meta-analysis was performed, describe the model(s), | Page 7-8 |

| Section and Topic | Item # | Checklist item | Location where item is reported |
|---|---|---|---|
| | | method(s) to identify the presence and extent of statistical heterogeneity, and software package(s) used. | |
| | 13e | Describe any methods used to explore possible causes of heterogeneity among study results (e.g. subgroup analysis, meta-regression). | Page 7-8 |
| | 13f | Describe any sensitivity analyses conducted to assess robustness of the synthesized results. | Page 7-8 |
| Reporting bias assessment | 14 | Describe any methods used to assess risk of bias due to missing results in a synthesis (arising from reporting biases). | Page 7 |
| Certainty assessment | 15 | Describe any methods used to assess certainty (or confidence) in the body of evidence for an outcome. | Page 7 |
| **RESULTS** | | | |
| Study selection | 16a | Describe the results of the search and selection process, from the number of records identified in the search to the number of studies included in the review, ideally using a flow diagram. | Page 8 |
| | 16b | Cite studies that might appear to meet the inclusion criteria, but which were excluded, and explain why they were excluded. | N/A |
| Study characteristics | 17 | Cite each included study and present its characteristics. | Page 8-12 |
| Risk of bias in studies | 18 | Present assessments of risk of bias for each included study. | Page 15<br>Supplementary Material |
| Results of individual studies | 19 | For all outcomes, present, for each study: (a) summary statistics for each group (where appropriate) and (b) an effect estimate and its precision (e.g. confidence/credible interval), ideally using structured tables or plots. | Page 16<br>Supplementary Material |
| Results of syntheses | 20a | For each synthesis, briefly summarise the characteristics and risk of bias among contributing studies. | Page 17-20 |
| | 20b | Present results of all statistical syntheses conducted. If meta-analysis was done, present for each the summary estimate and its precision (e.g. confidence/credible interval) and measures of statistical heterogeneity. If comparing groups, describe the direction of the effect. | Page 17-20 |
| | 20c | Present results of all investigations of possible causes of heterogeneity among study results. | Page 17-20<br>Supplementary Material |
| | 20d | Present results of all sensitivity analyses conducted to assess the robustness of the synthesized results. | Page 17-20<br>Supplementary Material |
| Reporting biases | 21 | Present assessments of risk of bias due to missing results (arising from reporting biases) for each synthesis assessed. | Page 15 |
| Certainty of evidence | 22 | Present assessments of certainty (or confidence) in the body of evidence for each outcome assessed. | Page 21-22 |
| **DISCUSSION** | | | |
| Discussion | 23a | Provide a general interpretation of the results in the context of other evidence. | Page 23-24 |
| | 23b | Discuss any limitations of the evidence included in the review. | Page 23-25 |
| | 23c | Discuss any limitations of the review processes used. | Page 25-26 |
| | 23d | Discuss implications of the results for practice, policy, and future research. | 27 |
| **OTHER INFORMATION** | | | |
| Registration and | 24a | Provide registration information for the review, including register name | N/A |

| Section and Topic | Item # | Checklist item | Location where item is reported |
|---|---|---|---|
| protocol | | and registration number, or state that the review was not registered. | |
| | 24b | Indicate where the review protocol can be accessed, or state that a protocol was not prepared. | N/A |
| | 24c | Describe and explain any amendments to information provided at registration or in the protocol. | N/A |
| Support | 25 | Describe sources of financial or non-financial support for the review, and the role of the funders or sponsors in the review. | Page 28 |
| Competing interests | 26 | Declare any competing interests of review authors. | Page 28 |
| Availability of data, code and other materials | 27 | Report which of the following are publicly available and where they can be found: template data collection forms; data extracted from included studies; data used for all analyses; analytic code; any other materials used in the review. | N/A |

*From:* Page MJ, McKenzie JE, Bossuyt PM, Boutron I, Hoffmann TC, Mulrow CD, et al. The PRISMA 2020 statement: an updated guideline for reporting systematic reviews. BMJ 2021;372:n71. doi: 10.1136/bmj.n71

**Supplemental Table 3: Risk of bias of the included studies - ROBINS-I tool results for non-randomized studies**

| Studies | Domains | | | | | | | |
|---|---|---|---|---|---|---|---|---|
| | Confusion | Selection | Classification | Deviations | Missing data | Measurement | Selection of the reported result | Overall bias |
| **Outcome: Mortality** | | | | | | | | |
| Wai, 2022 | Moderate | Low | Low | Moderate | Moderate | Low | Low | Moderate |
| Dryden-Peterson, 2022 | Low | Low | Low | Low | Moderate | Low | Low | Moderate |
| Wong, 2022 | Low | Low | Low | Low | Low | Low | Low | Low |
| Ganatra, 2022 | Low | Low | Low | Low | Low | Low | Low | Low |
| Aggarwal, 2022 | Low | Low | Serious | Low | Moderate | Low | Low | Serious |
| Najjar-Debbiny, 2022 | Low | Moderate | Low | Low | Low | Low | Low | Moderate |
| Bajema, 2022 | Low | Low | Low | Low | Moderate | Low | Low | Moderate |
| Patel, 2022 | Serious | Low | Low | Low | Moderate | Low | Low | Serious |
| Schwartz, 2023 | Low | Low | Low | Low | Moderate | Moderate | Low | Moderate |
| Hedvat, 2022 | Low | Moderate | Low | Low | Moderate | Moderate | Low | Moderate |
| Arbel, 2022 | Low | Low | Low | Low | Low | Low | Low | Low |
| Qian, 2022 | Low | Low | Low | Low | Moderate | Moderate | Low | Moderate |
| Yip, 2022 | Low | Low | Low | Low | Low | Low | Low | Low |
| **Outcome: Hospitalization** | | | | | | | | |
| Wai, 2022 | Moderate | Low | Low | Moderate | Moderate | Serious | Low | Serious |
| Dryden-Peterson, 2022 | Low | Low | Low | Low | Moderate | Serious | Low | Serious |
| Wong, 2022 | Low | Low | Low | Low | Low | Serious | Low | Serious |
| Ganatra, 2022 | Low | Low | Low | Low | Low | Serious | Low | Serious |
| Aggarwal, 2023 | Low | Low | Serious | Low | Serious | Serious | Low | Serious |
| Zhou, 2022 | Low | Serious | Low | Low | Low | Critical | Low | Critical |
| Patel, 2022 | Serious | Low | Low | Low | Moderate | Serious | Low | Serious |
| Arbel, 2022 | Low | Low | Low | Low | Low | Serious | Low | Serious |
| Yip, 2022 | Low | Low | Low | Low | Low | Moderate | Low | Moderate |
| Lewnard, 2022 | Low | Low | Low | Low | Low | Serious | Low | Serious |
| Shan, 2022 | Low | Low | Low | Low | Moderate | Moderate | Low | Moderate |

### Supplemental Table 4 - Results of effectiveness of the studies included in the review.

| Studies | *Time-point* | Alternative | Patients with event (N) | Total of patients (N) | % with event | Risk (95%CI) | p-value |
|---|---|---|---|---|---|---|---|
| **Mortality** | | | | | | | |
| Ganatra et al., 2022[a] | 30 days | Nirmatrelvir-ritonavir | 0 | 1,130 | 0 | RD: −0,009 (−0,014 a −0,003) | 0.002 |
| | | No antiviral treatment | 10 | 1,130 | 0.88 | | |
| Wai et al., 2022 | 30 days | Nirmatrelvir-ritonavir | 0 | 4442 | 0 | NR | NR |
| | | No antiviral treatment | 65 | 23,430 | 0.03 | | |
| Dryden-Peterson et al, 2022[a.e] | 28 days | Nirmatrelvir-ritonavir | 8 | 11,797 | 0.07 | aRR: 0.29 (0.12 a 0.71) | NR |
| | | No antiviral treatment | 62 | 32,248 | 0.19 | | |
| Hedvat et al., 2022 | 30 days | Nirmatrelvir-ritonavir | 0 | 28 | 0 | NR | NR |
| | | No antiviral treatment | 3 | 75 | 0.04 | NR | |
| Wong et al., 2022[a] | 28 days | Nirmatrelvir-ritonavir | 22 | 5,542 | 0.16 | HR: 0.34 (0.22 a 0.52) | < 0,0001 |
| | | No antiviral treatment | 635 | 54,672 | 0.68 | | |
| Arbel et al., 2022 | 35 days | Nirmatrelvir-ritonavir | 3 | 3,902 | 0.08 | NR | NR |
| | | No antiviral treatment | 174 | 105,352 | 0.2 | | |
| Bajema et al., 2022[a] | 30 días 31–180 days | Nirmatrelvir-ritonavir | 5 | 1,587 | 0.32 | RR: 0.21 (0.09-0.52) | NR |
| | | No antiviral treatment | 24 | 1,587 | 1.51 | | |
| Patel et al., 2022 | 28 days | Nirmatrelvir-ritonavir | 1 | 337 | 0.3 | NR | NR |
| | | No antiviral treatment | 75 | 4.044 | 1.85 | | |
| Zhou et al., 2022[a] | 30 days | Nirmatrelvir-ritonavir | 7 | 2,808 | 0.25 | 0.16 (0.11, 0.22) | NR |
| | | No antiviral treatment | 100 | 10,849 | 0.92 | | |
| Schwartz et. al., 2022 [a,d] | 30 days | Nirmatrelvir-ritonavir | 142 | 8,876 | 1.6 | OR: 0.49 (0.40 – 0.60) | |
| | | No antiviral treatment | 5,566 | 168,669 | 3.3 | | |
| Aggarwal et al., 2023[a] | 28 days | Nirmatrelvir-ritonavir | 2 | 7,168 | 0.03 | aOR: 0.15 (0.03-0.50) | 0.001 |
| | | No antiviral treatment | 15 | 9,361 | 0.2 | | |
| Shah et al., 2022[c] | 30 days | Nirmatrelvir-ritonavir | 29 | 198,927 | 0.01 | NR | NR |
| | | No antiviral treatment | 182 | 500,921 | 0.04 | | |
| Qian et al., 2022 | 30 days | Nirmatrelvir-ritonavir | 1 | 307 | 0.3 | NR | NR |
| | | No antiviral treatment | 2 | 278 | 0.7 | | |
| **Hospitalization** | | | | | | | |
| Ganatra et al., 2022 | 30 days | Nirmatrelvir-ritonavir | 10 | 1,130 | 0.88 | 0.43 (0.2-0.9) | 0.02 |
| | | No antiviral treatment | 23 | 1,130 | 2.04 | | |
| Yip et al., 2022[a] | 30 days | Nirmatrelvir-ritonavir | 177 | 4,921 | 3.6 | WHR: 0.79 (0.65–0.95) | 0.011 |
| | | No antiviral treatment | 214 | 4,758 | 4.5 | | |

| | | | | | | | |
|---|---|---|---|---|---|---|---|
| Dryden-Peterson et al, 2022[a,e] | 14 days | Nirmatrelvir-ritonavir | 61 | 11,797 | 0.52 | aRR: 0.60 (0.44 a 0.81) | NR |
| | | No antiviral treatment | 248 | 32,248 | 0.77 | | |
| Wai et al., 2022 | 30 days | Nirmatrelvir-ritonavir | NI | 4,442 | NI | OR: 0.37 (0.23, 0.60) | <0.0001 |
| | | No antiviral treatment | NI | 23,430 | NI | | |
| Wong et al., 2022[a] | 28 days | Nirmatrelvir-ritonavir | 246 | 5,542 | 4.29 | HR: 0.76 (0.67 a 0.86) | < 0.0001 |
| | | No antiviral treatment | 3185 | 54,672 | 5.62 | | |
| Arbel et al., 2022 | 35 days | Nirmatrelvir-ritonavir | 18 | 3,902 | 0.46 | NR | NR |
| | | No antiviral treatment | 1,093 | 105,352 | 1.04 | | |
| Bajema et al., 2022[a] | 30 days | Nirmatrelvir-ritonavir | 43 | 1,587 | 2.71 | RR: 0.66 (0.48-0.91) | NR |
| | | No antiviral treatment | 65 | 1,587 | 4.10 | | |
| Lewnard et. al., 2022[a] | 30 days | Nirmatrelvir-ritonavir | 46 | 7,274 | 0.6 | NR | NR |
| | | No antiviral treatment | 641 | 126,152 | 0.5 | | |
| Patel et al., 2022 | 30 days | Nirmatrelvir-ritonavir | 5 | 337 | 1.5 | NR | NR |
| | | No antiviral treatment | 251 | 4044 | 6.2 | | |
| Aggarwal et al., 2022[a] | 28 days | Nirmatrelvir-ritonavir | 61 | 7,168 | 0.9 | aOR: 0.45 (0.33-0.62) | <0.001 |
| | | No antiviral treatment | 135 | 9,361 | 1.4 | | |
| Zhou et al., 2022[a] | 30 days | Nirmatrelvir-ritonavir | 34 | 2,808 | 1.21 | HR: 0.16 (0.11, 0.22) | NR |
| | | No antiviral treatment | 752 | 10,849 | 6.94 | | |
| Shah et al., 2022 | 30 days | Nirmatrelvir-ritonavir | 930 | 198,927 | 0.47 | aHR: 0.49 (0.46-0.53) | NR |
| | | No antiviral treatment | 4.299 | 500,921 | 0.86 | | |
| Qian et al., 2022 | 30 days | Nirmatrelvir-ritonavir | 4 | 307 | 1.3 | NR | NR |
| | | No antiviral treatment | 49 | 278 | 17.6 | | |
| **Composite outcome of mortality, invasive mechanical ventilation, or ICU admission** | | | | | | | |
| Yip et al., 2022[a] | 30 days | Nirmatrelvir-ritonavir | 20 | 4,921 | 0.4 | aWHR: 0.81 (0.47–1.39) | 0.448 |
| | | No antiviral treatment | 24 | 4,758 | 0.5 | | |
| **Composite outcome of mortality or hospitalization** | | | | | | | |
| Hedvat et al., 2022 | 30 days | Nirmatrelvir-ritonavir | 4 | 28 | 14.3 | aRR: 0.21, (0.06 a 0.71) | 0.009 |
| | | No antiviral treatment | 25 | 75 | 33.3 | | |
| Dryden-Peterson et al, 2022 | 14 days and 28 days | Nirmatrelvir-ritonavir | 69 | 11,797 | 0.55 | aRR: 0.56, (0.42 a 0.75) | NR |
| | | No antiviral treatment | 310 | 32,248 | 0.97 | | |
| Bajema et al., 2022[a] | 30 days | Nirmatrelvir-ritonavir | 45 | 1,587 | 2.84 | OR: 0.53 (0.39-0.72) | NR |
| | | No antiviral treatment | 85 | 1,587 | 5.36 | | |
| Schwartz et. al., 2022[d] | 30 days | Nirmatrelvir-ritonavir | 187 | 8,876 | 2.1 | OR: 0.56 (0.47 – 0.67) | NR |
| | | No antiviral treatment | 6,241 | 168,669 | 3.7 | | |
| Lewnard et. al., 2022[a] | 30 days | Nirmatrelvir-ritonavir | 51 | 7,274 | 0.7 | Initiation of treatment < 5 | <0.01<br>0.03 |

| | | | | | | | |
|---|---|---|---|---|---|---|---|
| | | No antiviral treatment | 695 | 126,152 | 0.55 | days aHR: 0.20 (0.06, 0.66) Initiation of treatment > 5 days, aHR: 0.46 (0.23, 0.93) | |
| Qian et al., 2022 | 30 days | Nirmatrelvir-ritonavir | 4 | 307 | 1.3 | aOR: 0.12 (0.05, 0.25) | NR |
| | | No antiviral treatment | 49 | 278 | 17.6 | | |
| **Composite outcome of all-cause mortality, hospitalization, and emergency care** | | | | | | | |
| Ganatra et al., 2022[b] | 30 days | Nirmatrelvir-ritonavir | 89 | 1,130 | 7.87 | 0.50 (0.39, 0.67) | <0.001 |
| | | No antiviral treatment | 163 | 1,130 | 14.4 | | |
| Najjar-Debbiny et al., 2022[b] | 28 days | Nirmatrelvir-ritonavir | 39 | 4,737 | 0.82 | 0.54 (0.39–0.75) | <0.001 |
| | | No antiviral treatment | 903 | 175,614 | 0.51 | | |

aRR: adjusted Risk Ratio; RD: Risk difference; HR: Hazard Ratio; WHR: Weighted hazard ratio. a) Cohort matched by Propensity Score Matching (PSM) or by IPTW weighted analysis; b) Najjar-Debbiny et al., 2022, the composite outcome of disease worsening or mortality was considered in the all-cause mortality, hospitalization, and emergency care group. c) Wai et al., 2022 and Shah et. al., 2022 evaluated mortality in hospitalized patients; d) Data on deaths and hospital admission within 30 days of receiving nimatrelvir/ritonavir (n=8,876), or not (n=168,669), in weighted analysis using IPTW. e) Mortality and hospital admission data from the Dryden study -Peterson et al, 2022 were obtained by visual inspection of the cumulative incidence graph.

### Supplemental Fig 1: Forest plot of all-cause mortality outcome within 35 days - nirmatrelvir-ritonavir versus control - subgroups - published and unpublished studies.

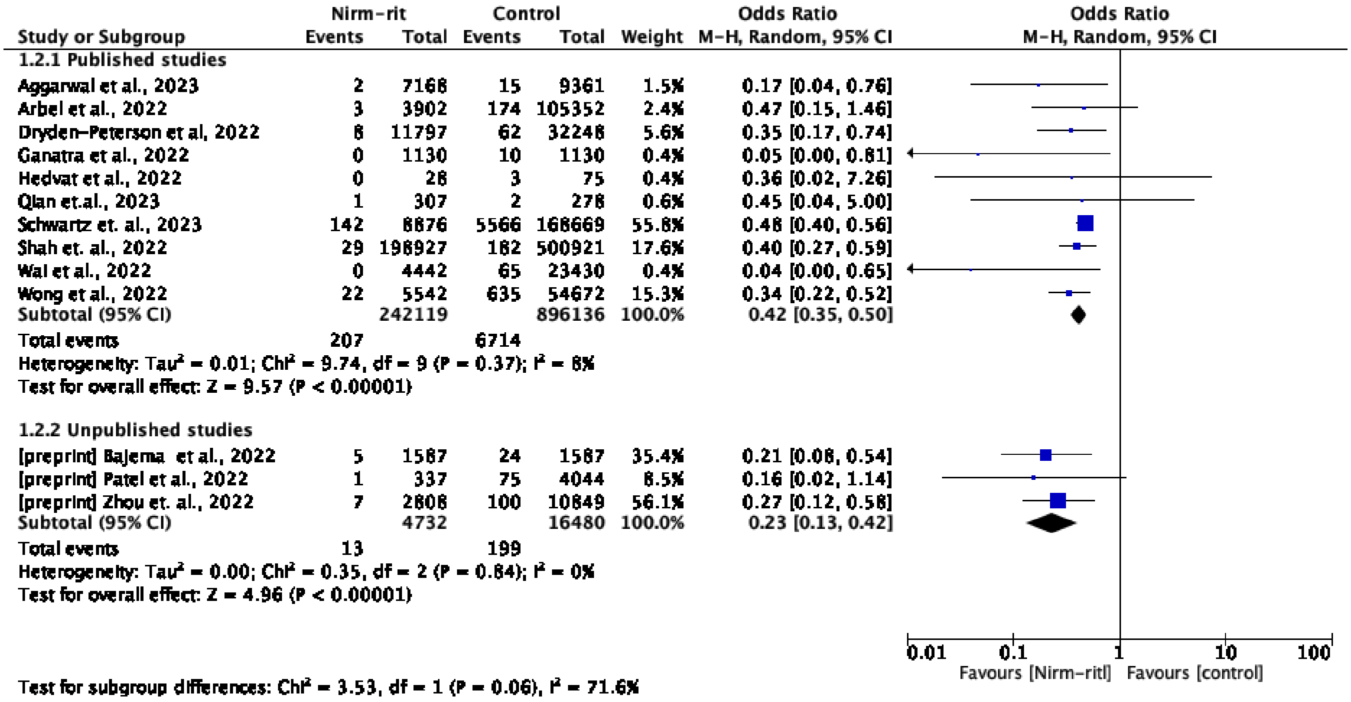

**Supplemental Fig 2: Forest plot of all-cause mortality outcome within 35 days - nirmatrelvir-ritonavir versus control - subgroups - matched and unmatched studies.**

| Study or Subgroup | Nirm-rit Events | Nirm-rit Total | Control Events | Control Total | Weight | Odds Ratio M-H, Random, 95% CI |
|---|---|---|---|---|---|---|
| **1.3.1 Unmatched studies** | | | | | | |
| [preprint] Patel et al., 2022 | 1 | 337 | 75 | 4044 | 3.2% | 0.16 [0.02, 1.14] |
| Arbel et al., 2022 | 3 | 3902 | 174 | 105352 | 9.7% | 0.47 [0.15, 1.46] |
| Hedvat et al., 2022 | 0 | 28 | 3 | 75 | 1.4% | 0.36 [0.02, 7.26] |
| Qian et.al., 2023 | 1 | 307 | 2 | 278 | 2.2% | 0.45 [0.04, 5.00] |
| Shah et. al., 2022 | 29 | 198927 | 182 | 500921 | 81.9% | 0.40 [0.27, 0.59] |
| Wai et al., 2022 | 0 | 4442 | 65 | 23430 | 1.6% | 0.04 [0.00, 0.65] |
| Subtotal (95% CI) | | 207943 | | 634100 | 100.0% | 0.38 [0.27, 0.54] |
| Total events | 34 | | 501 | | | |
| Heterogeneity: Tau² = 0.00; Chi² = 3.90, df = 5 (P = 0.56); I² = 0% | | | | | | |
| Test for overall effect: Z = 5.33 (P < 0.00001) | | | | | | |
| **1.3.3 Matched studies** | | | | | | |
| [preprint] Bajema et al., 2022 | 5 | 1587 | 24 | 1587 | 8.6% | 0.21 [0.08, 0.54] |
| [preprint] Zhou et. al., 2022 | 7 | 2808 | 100 | 10849 | 12.1% | 0.27 [0.12, 0.58] |
| Aggarwal et al., 2023 | 2 | 7168 | 15 | 9361 | 4.2% | 0.17 [0.04, 0.76] |
| Dryden-Peterson et al, 2022 | 8 | 11797 | 62 | 32248 | 12.8% | 0.35 [0.17, 0.74] |
| Ganatra et al., 2022 | 0 | 1130 | 10 | 1130 | 1.2% | 0.05 [0.00, 0.81] |
| Schwartz et. al., 2023 | 142 | 8876 | 5566 | 168669 | 37.3% | 0.48 [0.40, 0.56] |
| Wong et al., 2022 | 22 | 5542 | 635 | 54672 | 23.8% | 0.34 [0.22, 0.52] |
| Subtotal (95% CI) | | 38908 | | 278516 | 100.0% | 0.34 [0.25, 0.47] |
| Total events | 186 | | 6412 | | | |
| Heterogeneity: Tau² = 0.06; Chi² = 10.38, df = 6 (P = 0.11); I² = 42% | | | | | | |
| Test for overall effect: Z = 6.62 (P < 0.00001) | | | | | | |

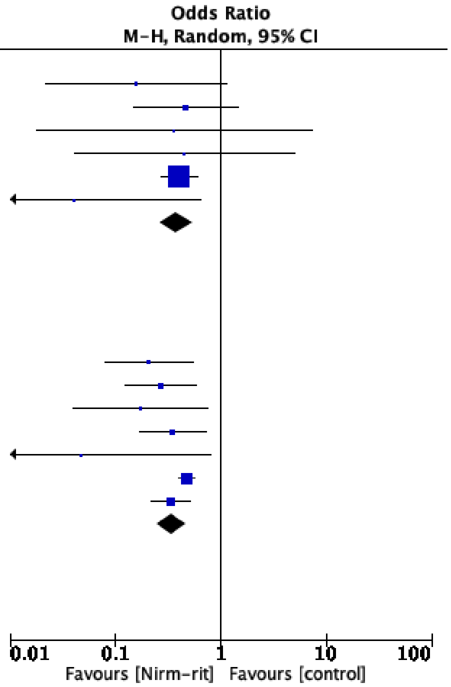


Test for subgroup differences: Chi² = 0.19, df = 1 (P = 0.66), I² = 0%

**Supplemental Fig 3: Forest plot of all-cause hospitalization outcome within 35 days - nirmatrelvir-ritonavir versus control - subgroups - published and unpublished studies.**

| Study or Subgroup | Nirm-rit Events | Nirm-rit Total | Control Events | Control Total | Weight | Odds Ratio M-H, Random, 95% CI |
|---|---|---|---|---|---|---|
| **1.7.1 Published studies** | | | | | | |
| Aggarwal et al., 2023 | 61 | 7168 | 135 | 9361 | 13.8% | 0.59 [0.43, 0.79] |
| Arbel et al., 2022 | 18 | 3902 | 1093 | 105352 | 10.0% | 0.44 [0.28, 0.71] |
| Dryden-Peterson et al, 2022 | 61 | 11797 | 248 | 32248 | 14.3% | 0.67 [0.51, 0.89] |
| Ganatra et al., 2022 | 10 | 1130 | 23 | 1130 | 5.7% | 0.43 [0.20, 0.91] |
| Qian et.al., 2023 | 4 | 307 | 49 | 278 | 3.5% | 0.06 [0.02, 0.17] |
| Shah et. al., 2022 | 930 | 198927 | 4299 | 500921 | 18.7% | 0.54 [0.51, 0.58] |
| Wong et al., 2022 | 238 | 5542 | 3075 | 54672 | 17.7% | 0.75 [0.66, 0.86] |
| Yip et al., 2022 | 177 | 4921 | 214 | 4758 | 16.3% | 0.79 [0.65, 0.97] |
| Subtotal (95% CI) | | 233694 | | 708720 | 100.0% | 0.57 [0.46, 0.71] |
| Total events | 1499 | | 9136 | | | |
| Heterogeneity: Tau² = 0.06; Chi² = 47.43, df = 7 (P < 0.00001); I² = 85% | | | | | | |
| Test for overall effect: Z = 5.13 (P < 0.00001) | | | | | | |
| **1.7.2 Unpublished studies** | | | | | | |
| [preprint] Bajema et al., 2022 | 43 | 1587 | 65 | 1587 | 35.2% | 0.65 [0.44, 0.96] |
| [preprint] Patel et al., 2022 | 5 | 337 | 251 | 4044 | 29.2% | 0.23 [0.09, 0.56] |
| [preprint] Zhou et. al., 2022 | 34 | 2808 | 752 | 10849 | 35.6% | 0.16 [0.12, 0.23] |
| Subtotal (95% CI) | | 4732 | | 16480 | 100.0% | 0.29 [0.10, 0.84] |
| Total events | 82 | | 1068 | | | |
| Heterogeneity: Tau² = 0.77; Chi² = 28.98, df = 2 (P < 0.00001); I² = 93% | | | | | | |
| Test for overall effect: Z = 2.29 (P = 0.02) | | | | | | |

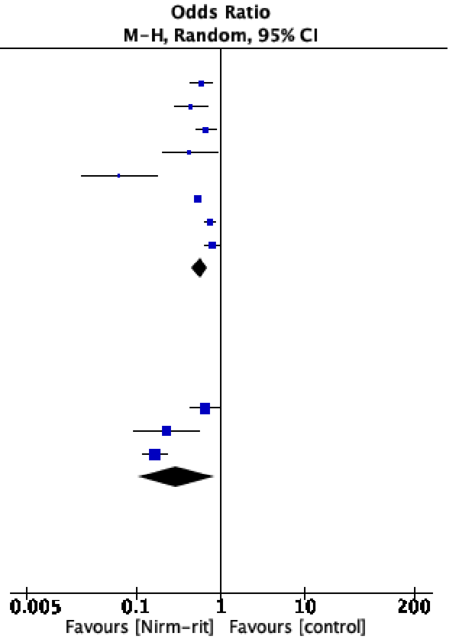


Test for subgroup differences: Chi² = 1.48, df = 1 (P = 0.22), I² = 32.3%

**Supplemental Fig 4: Forest plot of all-cause hospitalization outcome within 35 days - nirmatrelvir-ritonavir versus control - subgroups - matched and unmatched studies.**

| | Nirm-rit | | Control | | | Odds Ratio |
|---|---|---|---|---|---|---|
| Study or Subgroup | Events | Total | Events | Total | Weight | M-H, Random, 95% CI |
| 1.8.1 Unmatched studies | | | | | | |
| [preprint] Patel et al., 2022 | 5 | 337 | 251 | 4044 | 20.7% | 0.23 [0.09, 0.56] |
| Arbel et al., 2022 | 18 | 3902 | 1093 | 105352 | 28.2% | 0.44 [0.28, 0.71] |
| Qian et.al., 2023 | 4 | 307 | 49 | 278 | 18.4% | 0.06 [0.02, 0.17] |
| Shah et. al., 2022 | 930 | 198927 | 4299 | 500921 | 32.6% | 0.54 [0.51, 0.58] |
| Subtotal (95% CI) | | 203473 | | 610595 | 100.0% | 0.29 [0.15, 0.56] |
| Total events | 957 | | 5692 | | | |
| Heterogeneity: $Tau^2 = 0.36$; $Chi^2 = 21.18$, df = 3 (P < 0.0001); $I^2 = 86\%$ | | | | | | |
| Test for overall effect: Z = 3.65 (P = 0.0003) | | | | | | |
| 1.8.3 Matched studies | | | | | | |
| [preprint] Bajema et al., 2022 | 43 | 1587 | 65 | 1587 | 13.8% | 0.65 [0.44, 0.96] |
| [preprint] Zhou et. al., 2022 | 34 | 2808 | 752 | 10849 | 14.4% | 0.16 [0.12, 0.23] |
| Aggarwal et al., 2023 | 61 | 7168 | 135 | 9361 | 14.9% | 0.59 [0.43, 0.79] |
| Dryden-Peterson et al, 2022 | 69 | 11797 | 310 | 32248 | 15.3% | 0.61 [0.47, 0.79] |
| Ganatra et al., 2022 | 10 | 1130 | 23 | 1130 | 9.4% | 0.43 [0.20, 0.91] |
| Wong et al., 2022 | 238 | 5542 | 3075 | 54672 | 16.4% | 0.75 [0.66, 0.86] |
| Yip et al., 2022 | 177 | 4921 | 241 | 4758 | 15.9% | 0.70 [0.57, 0.85] |
| Subtotal (95% CI) | | 34953 | | 114605 | 100.0% | 0.52 [0.37, 0.73] |
| Total events | 632 | | 4601 | | | |
| Heterogeneity: $Tau^2 = 0.19$; $Chi^2 = 69.63$, df = 6 (P < 0.00001); $I^2 = 91\%$ | | | | | | |
| Test for overall effect: Z = 3.72 (P = 0.0002) | | | | | | |

Odds Ratio
M-H, Random, 95% CI

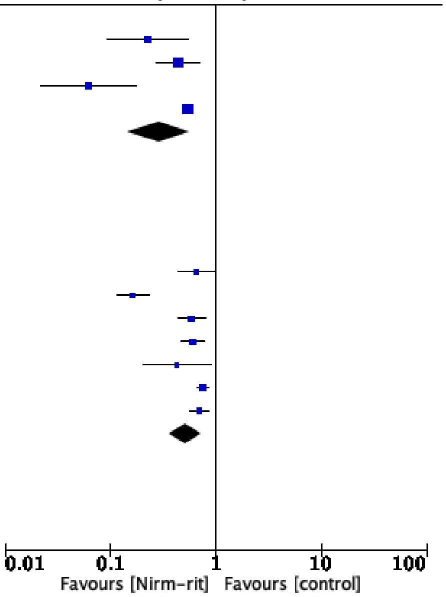


Test for subgroup differences: $Chi^2 = 2.37$, df = 1 (P = 0.12), $I^2 = 57.8\%$
